# An integrative eco-entomovirological study of *Culicoides* species associated with Oropouche virus in Colombia

**DOI:** 10.64898/2026.11.12.26367924

**Authors:** Juan S Mantilla-Granados, Santiago García-Guzmán, Cristian J Salcedo-Amórtegui, Eliana P Calvo, Daniela Ayos Cardozo, Jaime E. Castellanos

**Affiliations:** Universidad El Bosque, Vicerrectoría de Investigaciones, Grupo de Virología, Bogotá. 110121, Colombia; Secretaría de Salud del Vichada, Laboratorio Departamental de Salud Pública, Puerto Carreño, Vichada, Colombia

## Abstract

**Background:** Oropouche virus (OROV) causes recurrent outbreaks in South America, including Colombia. Although *Culicoides paraensis* is the principal recognized urban vector, the potential involvement of other *Culicoides* species remains incompletely understood. In Colombia, studies combining viral detection, host-feeding behavior, and ecological suitability are scarce.

**Methodology:** We investigated Culicoides communities in urban and peri-urban environments of Puerto Carreño, Vichada, during two seasonal transitions. Species identification combined morphology and COI DNA barcoding, and blood-meal sources were identified using vertebrate molecular markers. OROV RNA was assessed by RT- qPCR targeting the L and S genomic segments, and maximum likelihood prevalence estimates with 95% confidence intervals were calculated accounting for unequal pool sizes, including a conservative estimate based on concordant detection of both targets. Selected positive samples underwent multiplex amplicon sequencing fragments from all three OROV genomic segments (L, M, and S) using Oxford Nanopore Technology, followed by consensus reconstruction and phylogenetic analysis. Ecological suitability models were compared with reported OROV case distributions.

**Principal Findings:** We collected 1,639 specimens representing 11 taxa, eight supported by COI barcoding. *C. insignis* accounted for more than 70% of captures and was the only taxon collected in urban settings. OROV RNA was detected in *C. insignis*, *C. pusillus*, and *C. ignacioi*. In *C. insignis*, concordant detection of the L and S molecular targets yielded an estimated prevalence of 3.61 per 1,000 individuals (95% CI: 0.94–9.60). Nanopore sequencing recovered OROV-derived fragments spanning up to 656 nt across the L, M, and S genomic segments. Shorter fragments supported molecular identification, while phylogenetic analysis of a 656-nt M-segment region recovered from *C. insignis* and *C. pusillus* placed both sequences within a group containing viruses associated with the recent South American outbreak, although partial genomic coverage prevent formal lineage assignment. Blood-meal analyses identified feeding on humans, horses, and pigs in *C. insignis*; humans and horses in *C. pusillu*s; and horses in *C. ignacioi*. Ecological models identified distinct environmental associations among species, and predicted suitability for overlapping portions of regions reporting OROV cases. Administrative-scale concordance was heterogeneous.

**Conclusions:** The integration of molecular, host-feeding, and ecological evidence highlights *C. insignis*, *C. pusillus*, and *C. ignacioi* as species of epidemiological interest in the context of OROV circulation in Colombia. These findings expand current knowledge of OROV-associated *Culicoides* diversity and provide a basis for targeted entomological surveillance and experimental studies to clarify their potential roles in virus transmission.

**Author Summary:** Oropouche virus (OROV) causes recurrent outbreaks in South America, including Colombia. Although *Culicoides paraensis* is the principal recognized urban vector, the possible involvement of other biting midge species remains poorly understood. We investigated *Culicoides* communities in urban and peri-urban environments of Puerto Carreño, eastern Colombia, combining species identification, blood-meal analysis, viral RNA detection, sequencing, and ecological modeling. OROV RNA was detected by RT-qPCR in *C. insignis*, *C. pusillus*, and *C. ignacioi*. Genetic sequencing confirmed the presence of OROV-derived sequences. Partial viral M segment sequences grouped with references associated with recent South American outbreaks, although they did not allow formal lineage assignment. *C. insignis* was the most abundant species, with repeated viral detections and evidence of feeding humans and domestic animals. Ecological models showed distinct environmental associations among species and variable overlap with regions reporting human cases. Together, these findings highlight the potential epidemiological relevance of multiple *Culicoides* species and provide a basis for targeted surveillance and experimental studies to clarify their roles in OROV transmission.

## Introduction

Arboviruses pose a major public health threat as they can shift from zoonotic and sylvatic cycles to rural and urban transmission, driven by climate change, unplanned urbanization, human mobility, and land use change [1,2]. In tropical regions, environmental change and socioeconomic conditions can alter interactions among wildlife, vectors, and humans, creating opportunities for viral emergence and re-emergence [3]. The co-circulation of endemic arboviruses such as dengue, Zika, and chikungunya complicates clinical diagnosis and surveillance, contributing to the neglect of other emerging pathogens that may present with overlapping syndromes [4]. Historically, OROV has been associated with sylvatic cycles involving sloths as vertebrate hosts, while mosquitoes have been proposed as potential vectors in these cycles [5,6]. In urban settings in Brazil, the biting midge *Culicoides paraensis* is the principal vector[7]. However, in recent years, OROV has caused outbreaks across Brazil, Peru, Colombia, French Guiana, Panama, Ecuador, Venezuela, and Trinidad and Tobago, underscoring its regional expansion and growing clinical relevance [8]. While most infections manifest as self-limiting febrile syndromes, OROV has also been associated with meningitis, encephalitis, adverse pregnancy outcomes, and fatal cases, highlighting the need for an improved understanding of its transmission ecology and risk factors[1].

OROV, a member of the family *Peribunyaviridae* and genus *Orthobunyavirus*, possesses a tripartite negative-sense RNA genome with segments classified as small (S), medium (M), and large (L). The L segment encodes the RNA-dependent RNA polymerase, the M segment encodes glycoproteins involved in viral entry together with the non-structural protein NSm, and the S segment encodes the nucleoprotein and the interferon antagonist NSs [1]. This segmented genome allows genetic reassortment, an important mechanism of evolution in orthobunyaviruses [9]. Molecular assays targeting the L segment and the N gene within the S segment are used for OROV detection [10,11], while sequencing of viral genomic fragments can support identification and provide information on their relationships with reference strains.

Although *C. paraensis* has long been considered the primary urban vector, expanding geographic reports of OROV transmission raise the possibility that additional vector species may participate in local transmission cycles in distinct ecological contexts [12,13]. Establishing vector status requires complementary evidence from field and experimental studies, including accurate species identification, viral detection, host-feeding patterns, ecological associations, and experimental demonstration of vector competence[14,15]. Integrating these lines of evidence is particularly valuable for identifying candidate species and prioritizing them for subsequent experimental evaluation [14,16].

Although several mosquito species (family Culicidae) have been investigated as potential OROV vectors, *Culicoides paraensis*, a biting midge in the family Ceratopogonidae, remains the principal recognized urban vector. Experimental studies have also demonstrated susceptibility of *Culicoides sonorensis* to OROV under laboratory conditions [7,13,14]. *Culicoides* are hematophagous dipterans of the family Ceratopogonidae, whose minute size (<2 mm), cryptic species complexes, and wide distribution complicate their study. Molecular markers, such as the mitochondrial cytochrome c oxidase subunit I (COI) gene, aid species identification [17,18]. These midges feed on mammals, birds, reptiles, and amphibians [19,20] and act as potential bridges for zoonotic transmission. Their diverse breeding habitats, including muddy soils, marshes, riverbanks, manure, and decaying matter [21,22], make surveillance and control difficult. Given these biological characteristics, distinguishing the *Culicoides* species that may contribute to OROV transmission requires an approach that integrates taxonomy, viral detection and sequence characterization, and ecological context. Such integrative strategies are particularly important in regions where entomological data are sparse, and surveillance systems may underestimate viral circulation.

In Colombia, OROV circulation has been documented in the Amazon and border areas with Venezuela, During 2024–2025, 26 human OROV cases were included in the national epidemiological records compiled for this study, while the co-circulation of two viral lineages was documented in the Colombian Amazon in 2024 [8][23], but the *Culicoides* species potentially involved in local transmission remain poorly characterized. National research efforts on *Culicoides* have largely focused on their nuisance role, leaving major gaps in knowledge regarding species distribution, host-use patterns, and vector potential for arboviruses [24,25]. Vichada, with its mosaic of savannas, rainforests, and extensive hydrographic network, combined with socioeconomic dynamics linked to migration and land-use change [26,27], constitutes a strategic setting for investigating the contribution of *Culicoides* to OROV transmission. In this study, we aimed to characterize *Culicoides* species in urban and peri-urban areas of Puerto Carreño using an integrated approach combining morphological and molecular taxonomy, host-feeding analysis, and OROV RNA detection. We also evaluated species-specific ecological suitability and spatial concordance with reported OROV cases. Together, these analyses provide complementary evidence to prioritize candidate species for experimental vector competence studies and inform entomovirological surveillance of OROV in Colombia.

## Methods

### Study area

The study was conducted in the municipality of Puerto Carreño, the capital of the Vichada Department, Colombia, within the Orinoquía ecoregion at the confluence of the Meta and Orinoco rivers (Figure 1). This zone between the Orinoquía and Amazon Basin encompasses a complex mosaic of seasonally flooded savannas, *morichales* (palm-dominated wetlands), gallery forests, and temporary and permanent wetlands interconnected by an extensive hydrographic network [28]. Field collections were conducted under the institutional collection permit granted to Universidad El Bosque by the Autoridad Nacional de Licencias Ambientales (ANLA; Resolution No. 01470, November 17, 2017). The study protocol was approved by the Institutional Research Ethics Committee of Universidad El Bosque (Meeting Minutes No. 001-2024, January 23, 2024; Project Code CIE 2023-156).

**Figure 1.**
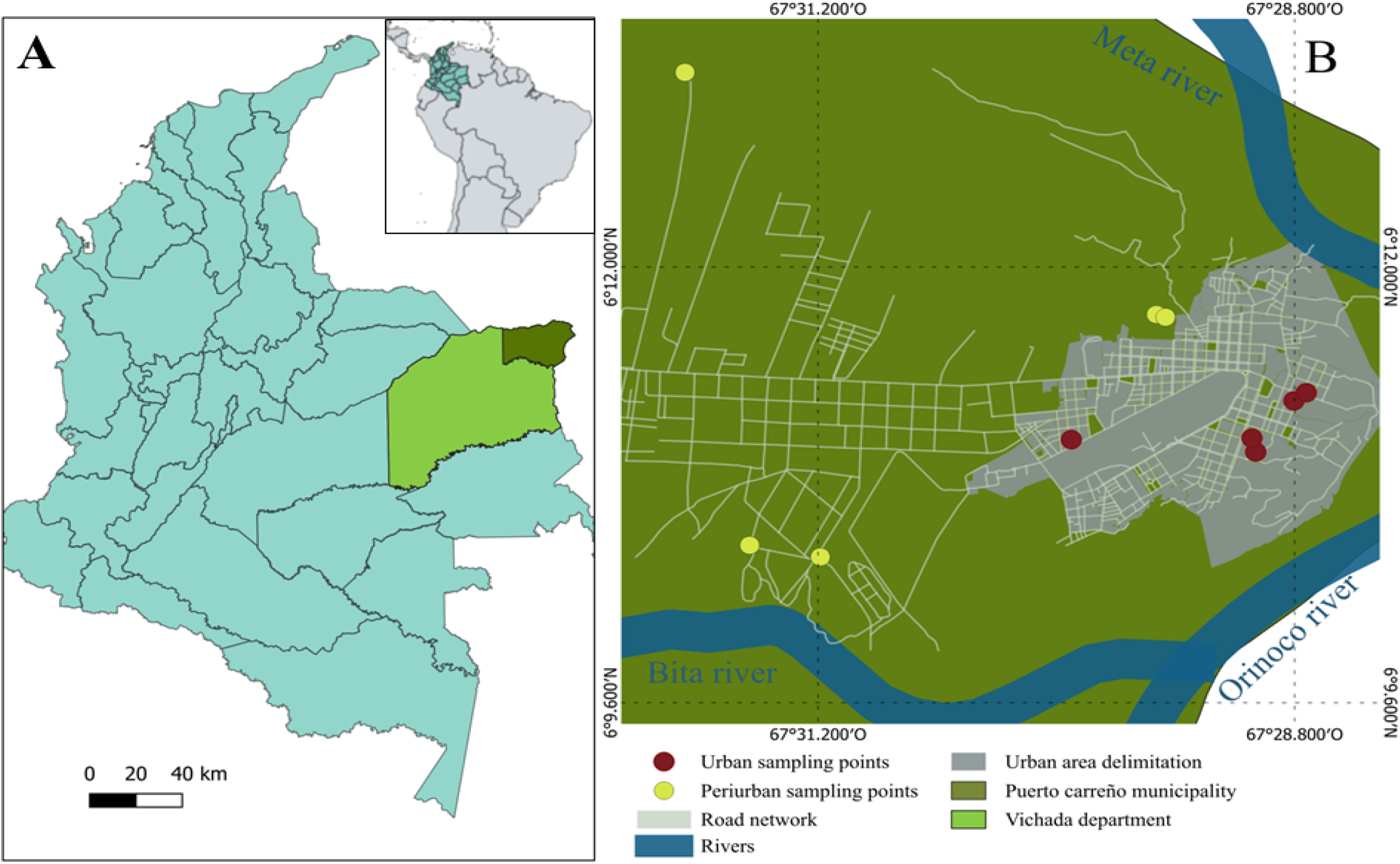
Sampling locations in Puerto Carreño, Colombia. The map shows the geographic position of the municipality and sampled areas. Administrative boundaries: DANE (Colombia), open-access data. Roads, rivers, and urban features: ©OpenStreetMap contributors, licensed under the Open Database License (ODbL).

### Entomological collections

Sampling sites were selected in urban and peri-urban areas of Puerto Carreño using environmental, entomological, and social criteria, including proximity to water bodies or secondary vegetation, the presence of domestic animals (pigs, chickens, and equids), community reports of biting nuisance, and official vector records from the Vichada Departmental Health Office. The final inclusion required the voluntary acceptance of the household owners.

Collections were conducted during two seasonal transition periods: rainy-to-dry (August–September 2024) and dry-to-rainy (May 2025). The sampling units corresponded to individual urban or rural households. At each site, two to five CDC ultraviolet light traps (model CDC 2506N, Curtis Dyna-Fog’s Ltd., USA) were deployed according to property size. Traps were placed both indoors and within 10 m of the dwelling, with placement determined by local land use and nearby habitats likely to host *Culicoides* breeding or aggregation sites, such as stagnant water organic matter accumulation (e.g., lagoons and banana crops when present), animal enclosures (pigsties, stables, chicken coops), and human living spaces.

The traps were operated for two consecutive nights at each site (17:00–08:00), and collection containers were retrieved each morning. Each trap-night was considered an independent sampling unit; therefore, a trap operated for two consecutive nights contributed two trap-nights. After each sampling round, traps were relocated to new households meeting the same selection criteria. When unusually high midge abundance was recorded at a specific location, additional collections were undertaken during the same fieldwork session. Capture abundance was standardized by the number of trap-nights to account descriptively for differences in trapping effort between sampling campaigns. Because sampling effort was unequal and collection sites were selected non-randomly, these standardized values were used to describe capture patterns rather than to infer seasonal changes in *Culicoides* abundance.

Sampling completeness was assessed through rarefaction analysis performed in R, using site-level species counts standardized by trap per night. Rarefaction curves enabled the estimation of species richness and evaluation of capture effort across sites, minimizing bias associated with unequal efforts.

### Field processing and specimen sorting

Specimens were transported to the Vichada Departmental Public Health Laboratory and chilled at −20 °C for approximately 2 h to induce temporary immobilization. Samples were sorted to retain *Culicoides* specimens, which were then separated into morphospecies using taxonomic keys [29–35]. Thus, individual morphological sorting preceded pooling, with representative specimens retained individually for taxonomic and molecular identification, blood-fed females separated for host-feeding analysis, and the remaining individuals pooled for viral RNA detection. Representative specimens of each morphospecies were preserved individually in RNAlater (Invitrogen) for nucleic acid stabilization. Blood-fed females were kept separately for blood meal source analysis, whereas the remaining individuals were grouped into pools of up to 60 specimens by species, collection date, and site for viral detection assays. All materials were shipped on dry ice to the Virology Group facilities at Universidad El Bosque (Bogotá, D.C., Colombia) and stored at −80 °C until downstream laboratory processing.

### Integrative taxonomy of *Culicoides* spp. Morphological determination

Representative specimens were processed using an integrative taxonomic approach to maintain morphological integrity and downstream suitability. Specimens were cleared in heated 85% lactic acid and dissected under glycerin. The mounts were examined using a Primo Star compound microscope and a Stemi 508 stereomicroscope (ZEISS), coupled to an Axiocam ERc 5s camera. Diagnostic structures, including the head, wings, and male or female terminalia, were mounted and examined in detail for species identification using dichotomous keys and regional monographs. Voucher specimens were preserved in glycerin-filled tubes as institutional reference material.

### DNA extraction and COI fragment amplification

Total DNA was extracted from individuals preserved in RNAlater using a commercial column-based kit (IBI Scientific, IB47222), according to manufacturer instructions. Prior to extraction, samples were mechanically homogenized with sterile plastic pestles in 1.5 mL microtubes containing DEPC-treated water, and then incubated for 2 h in the provided lysis buffer at room temperature for complete tissue digestion. DNA purity and yield were verified spectrophotometrically using a NanoPhotometer N120.

A 658 bp fragment of the COI gene was amplified using the universal primers (Table 1). Genomic DNA from *Aedes albopictus* C6/36 cells served as a positive control, and molecular-grade water was used as a non-template control. PCR products were verified by electrophoresis on 2% agarose gels stained with ethidium bromide. Samples showing visible bands were purified and sequenced by Sanger sequencing. The 30 COI sequences generated in this study were deposited in GenBank (Table S1).

**Table 1.**
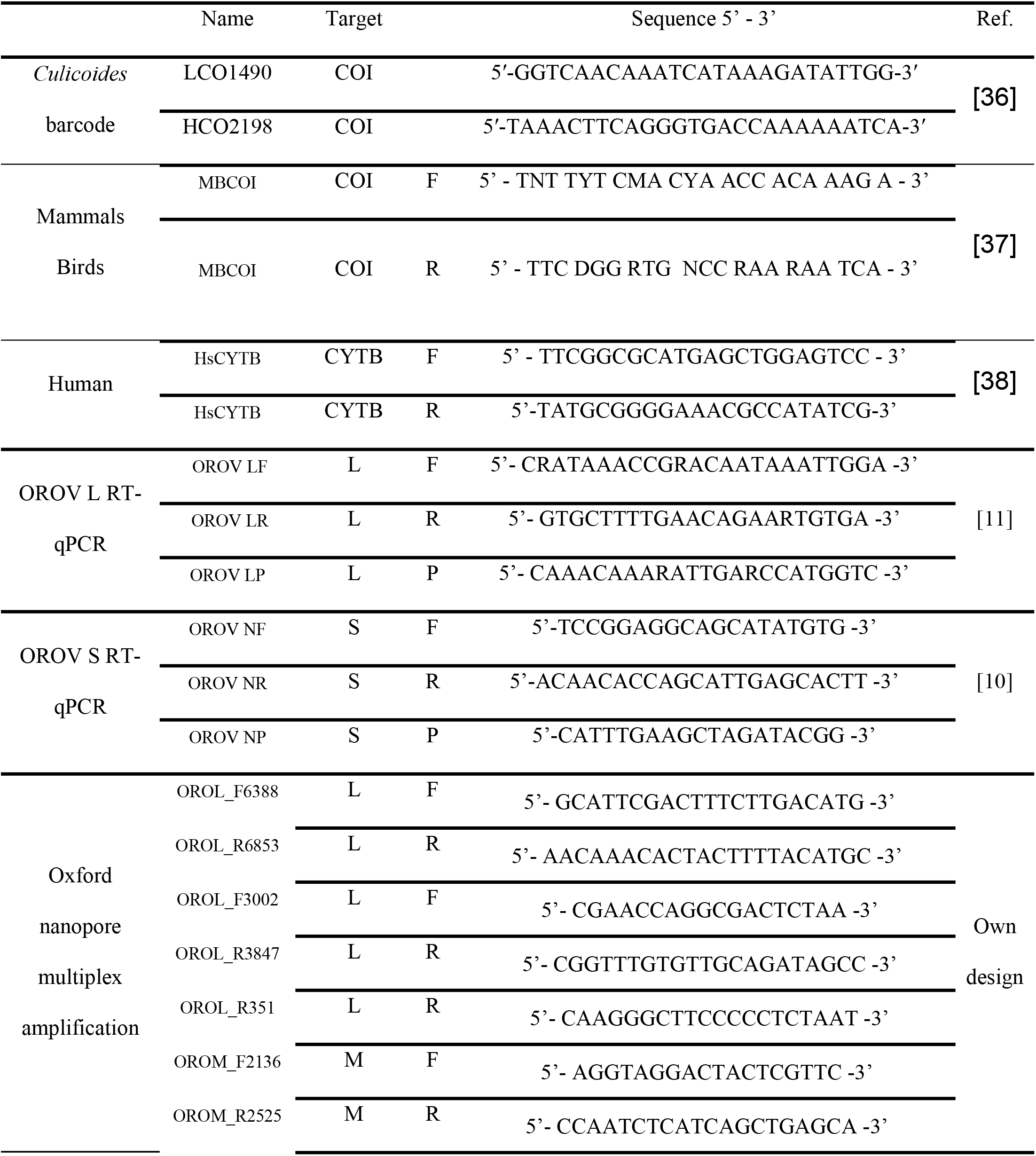
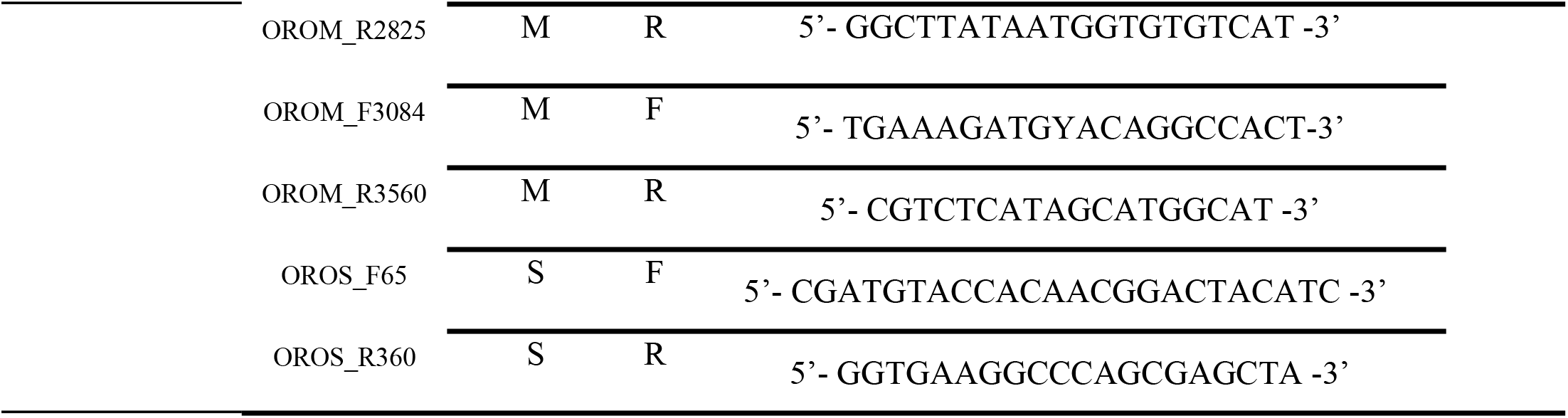
Primers used in this study.

### Barcode and phylogenetic analysis

Raw chromatograms were quality-filtered, trimmed, and assembled in DNAStar Lasergene® (SeqMan Pro). Forward and reverse reads were merged into consensus sequences with manual correction of ambiguous bases. Consensus sequences were aligned in MEGA v11 and compared with reference sequences of *Culicoides* or closely related Ceratopogonidae taxa retrieved from NCBI GenBank using BLAST, adopting a similarity threshold of >95% for species assignment.

Genetic distances were estimated using the Kimura two-parameter (K2P) model implemented in MEGA software. Phylogenetic relationships were inferred under a Bayesian framework using MrBayes v3.2.7 based on the multiple sequence alignment of newly generated sequences together with reference sequences retrieved from GenBank. Genetic distances were calculated primarily for comparative purposes and not used for phylogenetic inference. The best-fitting nucleotide substitution model (GTR+I+Γ) was selected using ModelFinder as implemented in IQ-TREE under the Bayesian Information Criterion (BIC). Two independent runs, each with four Markov chains (one cold and three heated), were performed for 2,000,000 generations, sampling every 100 generations. The first 25% of the samples were discarded as burn-in. Convergence was assessed by examining the effective sample sizes (ESS > 200), potential scale reduction factors (PSRF ≈ 1.0), and average standard deviation of split frequencies (ASDSF < 0.01). Posterior probabilities ≥ 0.95 were considered strong support for monophyletic relationships.

### Haplotype network analysis

The genetic diversity and spatial population structure of *Culicoides insignis*, the most abundant species in the collections and the one most frequently represented in the NCBI database, were examined. A haplotype network was constructed to visualize relationships among *C. insignis* COI haplotypes and their geographic distribution. Each haplotype is represented as a colored node, with connecting branches denoting single mutational steps. The COI sequences generated in this study were combined with georeferenced *C. insignis* sequences retrieved from GenBank®. Alignment was performed using MEGA v11, and the resulting dataset was exported in NEXUS/PAUP format. Haplotype definition was assigned by locality in DNA Sequence Polymorphism (DnaSP) v6.12.03 and PopArt v1.7 using the minimum spanning haplotype network algorithm. This approach enabled comparison of local haplotypes with those reported from other regions across the Neotropical range of *Culicoides insignis*.

### Blood-meal source analysis

The identification of vertebrate hosts was conducted on DNA extracted from blood-fed females using conventional PCR, following a previously published protocol [37] with modifications to the thermal cycling parameters to optimize the amplification efficiency. Gradient PCR assays were first calibrated with control blood samples from human, chicken, and duck hosts to define the optimal annealing conditions. Universal vertebrate primers and human-specific primers (Table 1) were used sequentially to detect single or mixed blood meal sources. Each reaction was performed in a final volume of 20 µL, containing 1× qScript PCR Kit (QuantaBio), 0.4 µM of forward and reverse primers, 1 µL of genomic DNA, and nuclease-free water. PCR products were resolved by electrophoresis on 2% agarose gels and documented using a gel imaging system. Extracted DNA from uninfected C6/36 *Aedes albopictus* cell line and from a *Culicoide*s male were used as negative controls, while nuclease-free water was used as a reaction control. Samples displaying amplicons of the expected size and sufficient intensity were purified and subjected to Sanger sequencing. The same samples were evaluated using both the universal vertebrate primers and the human-specific primers. The resulting chromatograms were edited in MEGA v12.0.11, and sequence identity was confirmed through BLASTn searches against the NCBI GenBank database. Host species were assigned based on a ≥ 98% sequence identity. This approach enabled the direct identification of vertebrate feeding sources and assessment of potential vector–host associations.

### OROV RNA detection analysis

Viral RNA detection was assessed in pools of female *Culicoides*. Pool size varied according to species, collection site, and sampling date, ranging from 2 to 60 individuals. Specimens were preserved immediately after collection in RNAlater to maintain RNA integrity under remote field conditions. RNA extracted from the pools was initially screened for OROV using two independent one-step RT-qPCR assays targeting the L and S genomic segments (Table 1). RNA from the same pools was not always available for both assays, resulting in different numbers of pools tested for the L and S targets. The L-segment assay used TaqMan chemistry and amplified an approximately 120-bp region, whereas detection of the S segment was performed using an LNA-based assay targeting an approximately 320-bp region. Reactions were performed using the qScript One-Step RT-qPCR Kit (QuantaBio) in a final volume of 15 µL, containing 1× qScript buffer, 0.4 µM of each primer, 0.4 µM probe, nuclease-free water, and 5 µL of RNA template. Appropriate negative extraction and no-template controls were included in each assay. Pools with cycle threshold (Ct) values below 37 were considered OROV-positive (Table S2). OROV RNA was extracted from infected cell culture supernatants denominated LET-01 and LET-05, in separate laboratory areas and biosafety cabinets from those used for the field-collected *Culicoides* samples. During RT-qPCR setup, positive-control templates were added last, after the sample tubes had been placed in the thermocycler. RNA extracted from uninfected C6/36 *Aedes albopictus* cells served as a negative extraction control, and nuclease-free water was included as a no-template control in each run.

### Pooled infection rate estimation

Because pool sizes varied among samples, OROV prevalence was estimated using a pooled-binomial maximum likelihood approach that explicitly accounts for individual pool sizes. Estimates and 95% confidence intervals were calculated using the pooledBin function in the R package binGroup (v2.2.3), with the maximum likelihood estimator and skew-score confidence intervals and are reported per 1,000 individuals. Estimates were calculated separately for the L and S molecular targets and, using a more conservative criterion, for pools showing concordant positivity for both targets. This criterion reflects the concurrent detection of two genomic segments in *Culicoides* species not previously characterized in this context.

### Nanopore sequencing and phylogenetic analysis of OROV

Samples were selected for amplicon sequencing using the Oxford Nanopore platform based on RNA availability and RT-qPCR results. Samples with Cq values below 36 for either target and samples positive for both the L and S targets were prioritized for sequencing. Multiplex amplification was performed using primer sets targeting L, M, and S (Table 1) genomic segments. Sequencing reads were filtered by length (250–800 bp) and mapped independently against OROV reference sequences using Minimap2 with settings optimized for Oxford Nanopore reads. Only primary alignments with a mapping quality (MAPQ) ≥20 were retained for downstream analyses. Read-derived consensus sequences were reconstructed independently for each sample and genomic region. Positions with a minimum read depth of 10× were retained, and the nucleotide supported by the 70% of reads at each position was incorporated into the consensus sequence; positions with equal support for alternative nucleotides were coded as ambiguous (N). Reference nucleotides were not used to fill uncovered or insufficiently supported positions, and discontinuous regions of coverage were retained as independent fragments. Negative controls were included during the Nanopore sequencing workflow to monitor potential contamination during library preparation and sequencing.

To assess sequence concordance and potential laboratory-source similarity, field-derived consensus sequences were compared with OROV control sequences generated independently by Sanger sequencing. Comparisons were restricted to homologous overlapping regions and included calculation of nucleotide identity and identification of nucleotide differences. For discrepant positions, nucleotide calls in the Nanopore consensus were evaluated against the underlying mapped-read data, including read depth, nucleotide counts, and majority-allele frequency. Pairwise genetic distances were calculated as uncorrected p-distances, representing the proportion of nucleotide differences across homologous positions, with gaps and ambiguous nucleotides excluded pairwise. Distances were calculated separately among field-derived sequences and between field-derived sequences and laboratory controls for the L (173 nt), M (316 nt), and S (299 nt) segments. For the extended M-segment dataset (656 nt), distances between B76/B82 and publicly available reference sequences were calculated and summarized according to previously assigned phylogenetic groups.

For phylogenetic analyses, Nanopore-derived consensus sequences were aligned with publicly available OROV sequences representing the genetic diversity of the corresponding genomic segments from GISAID up to September 30, 2026. Alignments were constructed separately for the L, M, and S segments and restricted to homologous genomic regions; non-overlapping fragments were not concatenated. The 13 field-derived sequences were initially aligned separately for each genomic segment. For phylogenetic analyses, sequence subsets were selected according to fragment length and the availability of homologous regions in the reference sequences, aiming to retain the largest possible number of samples while maintaining a sufficiently long region for comparison. Because fragment lengths varied among samples, separate alignments were generated for some segments. For the M segment, two alignments covering the same genomic region but differing in sequence length and sample inclusion were analyzed. Most S-segment fragments were approximately 90 bp long and were excluded from phylogenetic analysis when their limited length did not allow meaningful comparison with reference sequences.

Nucleotide substitution models were evaluated using IQ-TREE 2, and Bayesian phylogenetic inference was performed in MrBayes using the selected segment-specific models. Markov chain Monte Carlo analyses were run for 5 × 10⁶ generations, with convergence evaluated using the average standard deviation of split frequencies, effective sample sizes, and potential scale reduction factors. Node support was expressed as posterior probabilities. Phylogenetic placement of field-derived sequences was evaluated in the context of publicly available OROV sequences and previously reported phylogenetic group assignments.

### Ecological niche modeling

To infer the potential geographic distributions of the *Culicoides* species detected in this study, ecological niche models were generated using occurrence records from field collections supplemented with georeferenced data from the Global Biodiversity Information Facility (GBIF; https://www.gbif.org) and previously published datasets [25,39–46]. Records were curated to remove duplicates, spatial outliers, and entries that lacked precise geographic coordinates. Only georeferenced records with adequate spatial resolution were retained. Because the objective was to compare species-specific environmental structuring at a continental scale rather than to produce fine-resolution predictive maps, occurrence records were subjected to quality control before ecological niche modeling. Records with missing or invalid geographic coordinates, exact duplicate coordinate pairs, and locations outside the predefined geographic extent of the Americas (120°W–30°W; 60°S–35°N) were excluded. Occurrences with missing values for the environmental predictors retained in each species-specific model were also removed. No minimum-distance spatial thinning was applied; therefore, geographically proximate records with distinct coordinates could remain in the final occurrence datasets.

Environmental predictors representing climatic, topographic, vegetational, and edaphic attributes were obtained from publicly available global datasets. Bioclimatic variables (BIO1–BIO19), representing long-term summaries of temperature and precipitation, were downloaded from WorldClim v2.1 (1970–2000 normals; https://www.worldclim.org) at 30 arc-second (∼1 km) resolution. The mean annual wind speed at a height of 10 m was derived from the WorldClim monthly climate layers. Elevation data were obtained from NASADEM_HGT/001 (https://lpdaac.usgs.gov), and the terrain slope was computed using QGIS. Vegetation productivity was characterized using long-term MODIS MOD13 products (https://lpdaac.usgs.gov), and soil conditions were represented by the soil organic carbon content in the upper 0–5 cm layer from SoilGrids (https://soilgrids.org). All raster layers were resampled and harmonized to a spatial resolution of ∼ 1 km in WGS84 (EPSG:4326) and clipped to a continental extent encompassing the documented biogeographic distribution of the studied taxa across the Americas (120°W to 30°W; 60°S to 35°N) prior to export from Google Earth Engine.

Species distribution models were implemented in R (v4.5.1) using the maximum entropy algorithm (Maxnet package) and spatial raster tools from the terra package. Multicollinearity among environmental predictors was assessed independently for each species using the variance inflation factor (VIF). A threshold of VIF ≤ 10 was applied to identify and exclude highly collinear predictors, following established methodological criteria commonly used in ecological modeling and species distribution studies. For each species, a target of 2,000 background points was specified within the modeling extent. However, the effective number of background points used in model fitting varied among species, as determined by the species-specific modeling datasets and the availability of environmental information. The final number of background points used for each species is reported in Table 5. For each species, occurrence records were randomly partitioned into training (70%) and testing (30%) subsets, without spatial stratification. Habitat suitability surfaces were projected using a cloglog transformation. The modeling extent was defined to encompass the maximum documented geographic distributions reported for the studied taxa in previous systematic and faunistic studies, particularly for *Culicoides insignis*, thereby avoiding extrapolation beyond its known continental range.

Model performance was assessed using the area under the receiver operating characteristic curve (AUC) and the True Skill Statistic (TSS), calculated on independent test data (pROC package). Binary suitability maps were derived using thresholds that maximized the sensitivity and specificity (Youden’s index). Variable importance was quantified using a drop-AUC approach, in which each retained predictor was sequentially removed, and the resulting reduction in AUC in the test data was recorded. Predictors yielding positive drop-AUC values were interpreted as having independent explanatory contributions (“effective predictors”).

To evaluate interspecific ecological similarity, pairwise Jaccard similarity indices were calculated based on the presence or absence of effective predictors for each species in the study. This metric quantifies shared, independently informative environmental drivers rather than spatial niche overlap. Final suitability maps were generated for *Culicoides insignis*, *C. pusillus*, *C. ignacioi*, and *C. paraensis* to compare the ecological structuring and potential geographic overlap among potential Oropouche virus epidemiological interest *Culicoides* species.

### Species distribution and OROV case overlay

To assess the spatial concordance between predicted *Culicoides* species suitability and reported human OROV cases, species-specific suitability maps were overlaid with reported case distributions to visualize spatial co-occurrence patterns across the Americas. A fixed suitability threshold (≥ 0.4) was used to define high-suitability areas. This value approximated the range of species-specific optimal discrimination thresholds derived from model performance metrics and was consistently applied across taxa to enable standardized interspecific comparisons. The suitable areas were derived from Maxent models and projected as continuous cloglog outputs. Reported human OROV case data were compiled from the Pan American Health Organization (PAHO/WHO) and national surveillance reports from Brazil (11,888 cases), Panama (501), Peru (330), Colombia (26), Cuba (28), Venezuela (5), and Guyana (1) [8,11]. For all countries except Venezuela (where data were available only at the national level), cases were assigned to first-order administrative units (e.g., departments or provinces).

To formally evaluate administrative-scale ecological concordance, suitability values were summarized within each first-order administrative unit by calculating the mean suitability. Differences between regions with and without reported OROV cases were assessed using Wilcoxon rank-sum test. In addition, suitability values were dichotomized using a fixed threshold (≥ 0.4) to generate binary high/low suitability categories, which were cross-tabulated against case presence to compute odds ratios (OR) and 95% confidence intervals using Fisher’s exact test.

## Results

A total of 1,639 *Culicoides* specimens were collected across the two sampling campaigns conducted in Puerto Carreño, Vichada. Of these, 1,618 individuals were captured during the transition from rainy-to-dry weather, whereas 21 were collected during the transition from dry-to-rainy weather. Specimens were assigned to 11 morphologically defined taxa. The rarefaction curve (Figure S1) suggested that sampling completeness was approaching an asymptote.

As summarized in Table 2, relative abundance differed markedly between urban and peri-urban settings and between sampling periods. During the transition from rainy-to-dry, 99.8% of specimens were captured in peri-urban sites and only 0.2% in urban areas. All the specimens collected in urban areas were identified as *Culicoides insignis*. This species was the dominant in all the collections, representing more than 70% of the collected specimens.

**Table 2.**
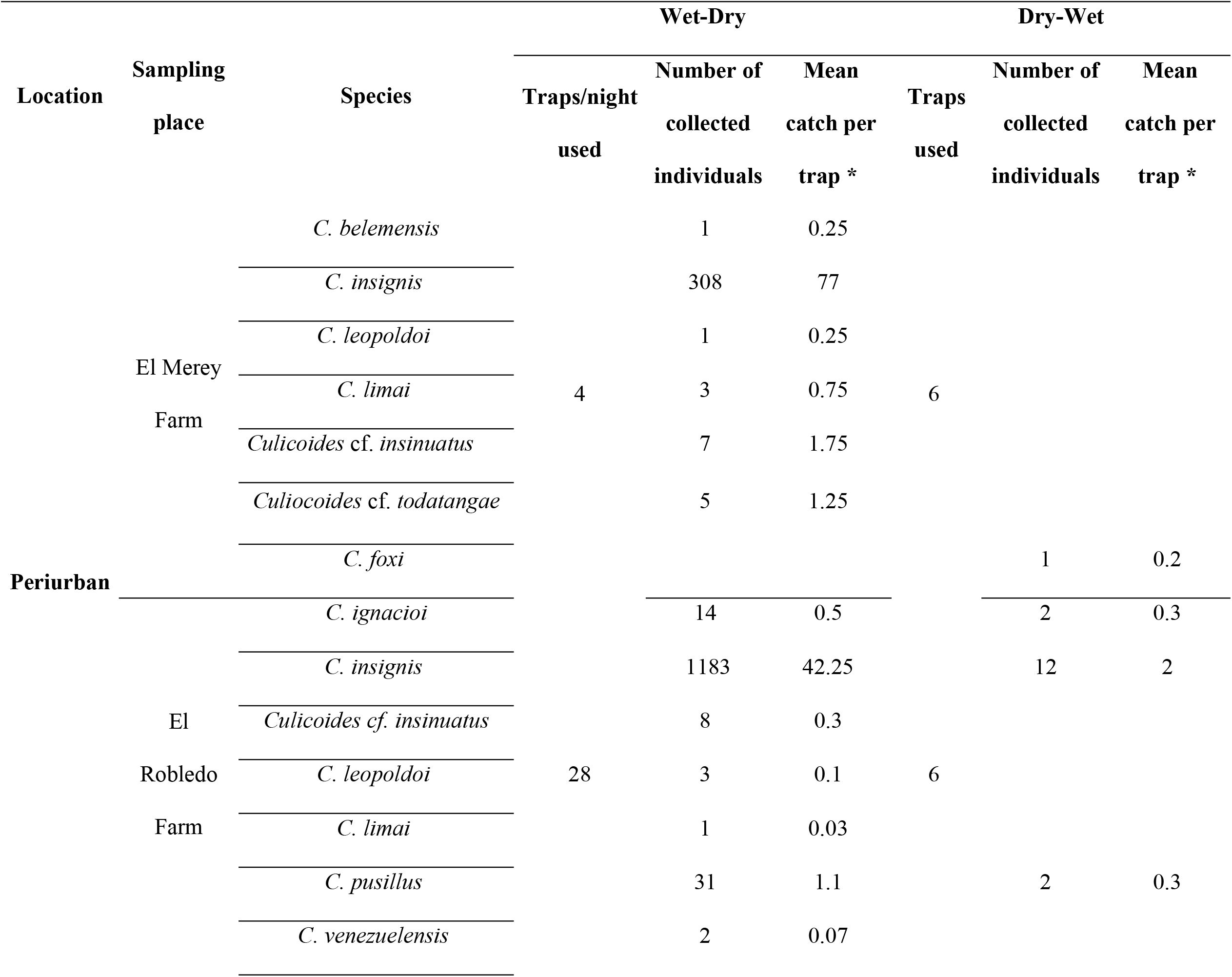
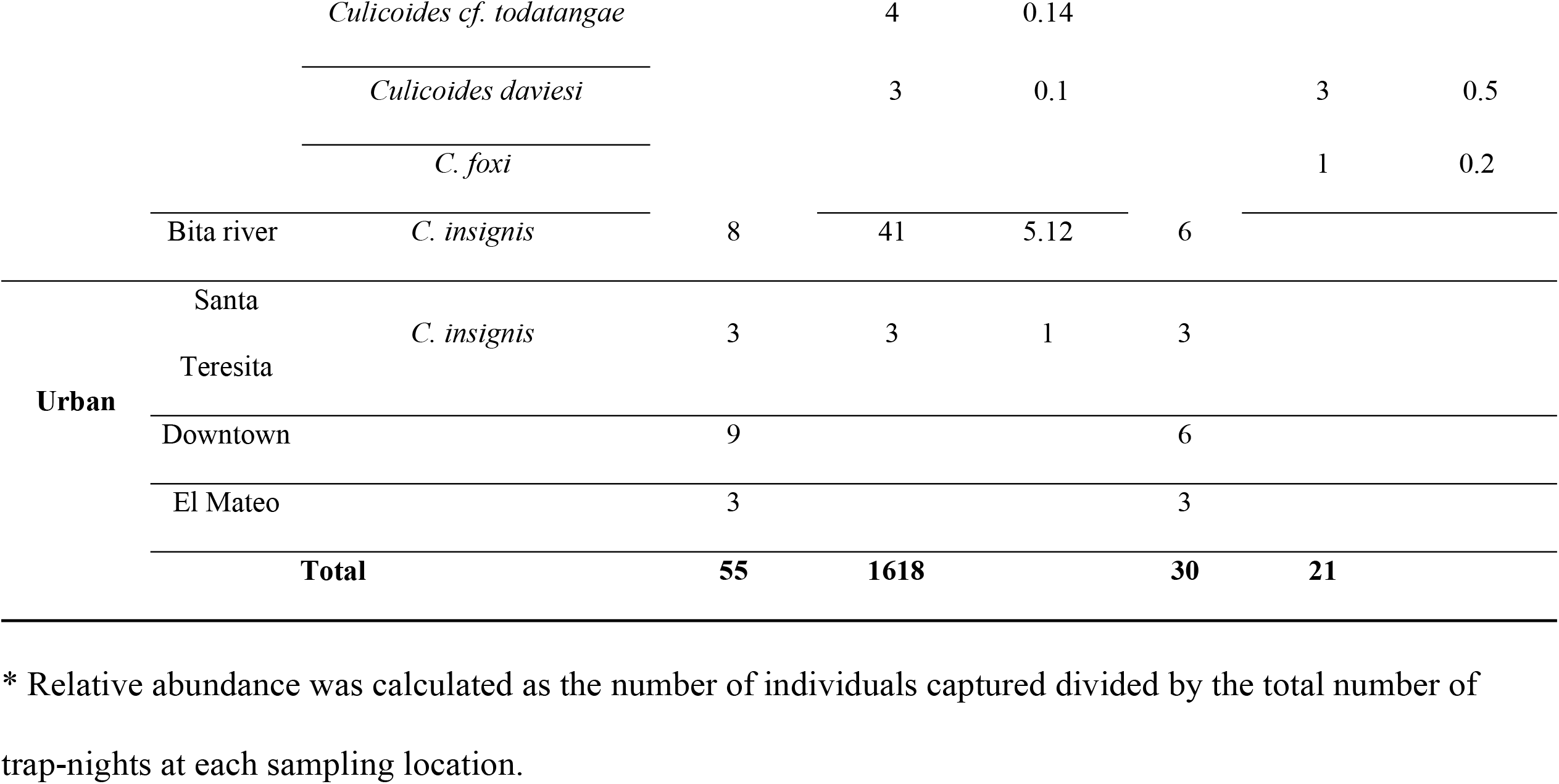
Species records and Mean catch per trap of *Culicoides* collected in Puerto Carreño, Vichada, across two sampling periods in periurban and urban areas.

|  |  |  | **Wet-Dry** |  |  | **Dry-Wet** |  |  |
|---|---|---|---|---|---|---|---|---|
| **Location** | **Sampling place** | **Species** | **Traps/night used** | **Number of collected individuals** | **Mean catch per trap \*** | **Traps used** | **Number of collected individuals** | **Mean catch per trap \*** |
| **Periurban** | El Merey Farm | *C. belemensis* | 4 | 1 | 0.25 | 6 |  |  |
|  |  | *C. insignis* |  | 308 | 77 |  |  |  |
|  |  | *C. leopoldoi* |  | 1 | 0.25 |  |  |  |
|  |  | *C. limai* |  | 3 | 0.75 |  |  |  |
|  |  | *Culicoides* cf. *insinuatus* |  | 7 | 1.75 |  |  |  |
|  |  | *Culiocoides* cf. *todatangae* |  | 5 | 1.25 |  |  |  |
|  |  | *C. foxi* |  |  |  |  | 1 | 0.2 |
|  | El Robledo Farm | *C. ignacioi* | 28 | 14 | 0.5 | 6 | 2 | 0.3 |
|  |  | *C. insignis* |  | 1183 | 42.25 |  | 12 | 2 |
|  |  | *Culicoides cf. insinuatus* |  | 8 | 0.3 |  |  |  |
|  |  | *C. leopoldoi* |  | 3 | 0.1 |  |  |  |
|  |  | *C. limai* |  | 1 | 0.03 |  |  |  |
|  |  | *C. pusillus* |  | 31 | 1.1 |  | 2 | 0.3 |
|  |  | *C. venezuelensis* |  | 2 | 0.07 |  |  |  |
|  |  | *Culicoides cf. todatangae* |  | 4 | 0.14 |  |  |  |
|  |  | *Culicoides daviesi* |  | 3 | 0.1 |  | 3 | 0.5 |
|  |  | *C. foxi* |  |  |  |  | 1 | 0.2 |
|  | Bita river | *C. insignis* | 8 | 41 | 5.12 | 6 |  |  |
| **Urban** | Santa Teresita | *C. insignis* | 3 | 3 | 1 | 3 |  |  |
|  | Downtown |  | 9 |  |  | 6 |  |  |
|  | El Mateo |  | 3 |  |  | 3 |  |  |
|  | **Total** |  | **55** | **1618** |  | **30** | **21** |  |

\* Relative abundance was calculated as the number of individuals captured divided by the total number of trap-nights at each sampling location.

### Species barcode analysis

Phylogenetic analysis of the mitochondrial COI gene supported the morphological identification of several *Culicoides* taxa. Sequences clustered into well-supported clades (posterior probabilities ≥ 0.95; Figure 2), generally corresponding to recognized subgenera and morphologically assigned species. Specimens identified as *Culicoides insignis* formed a strongly supported clade together with reference sequences retrieved from GenBank. Within this clade, internal sub structuring was observed, consistent with intraspecific haplotype diversity.

**Figure 2.**
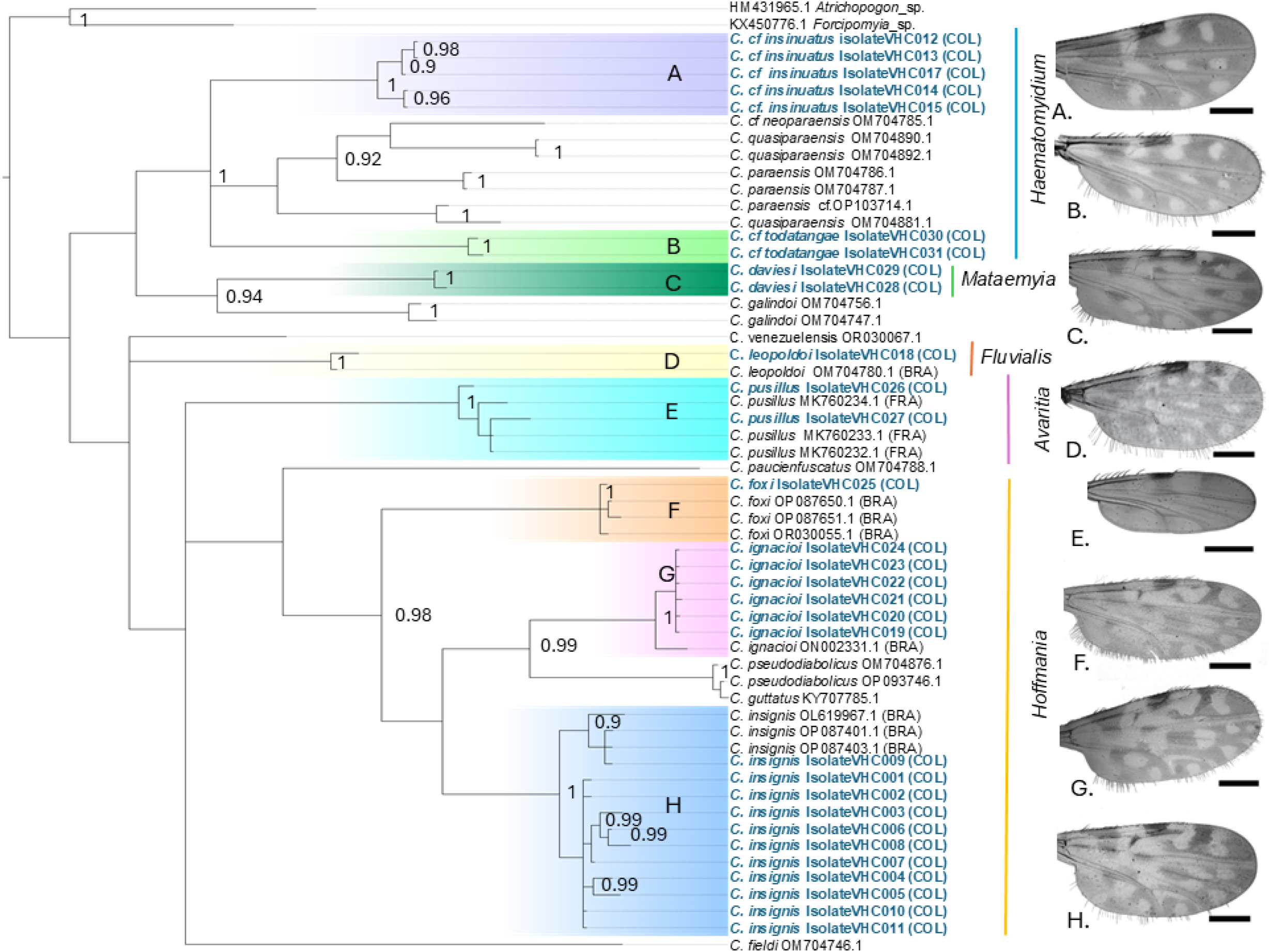
Bayesian phylogenetic reconstruction of *Culicoides* COI sequences and concordance with morphological identification. Bayesian inference was conducted on a 358-bp fragment of the mitochondrial cytochrome c oxidase subunit I (COI) gene under the GTR+I+Γ substitution model selected using ModelFinder (BIC criterion). Posterior probabilities ≥ 0.90 are indicated at nodes. Clades corresponding to morphologically delimited taxa are highlighted in distinct colors and labeled (A–H). Representative wing pigmentation patterns used for species diagnosis are shown on the right to illustrate concordance between molecular clustering and diagnostic morphology. Sequences of *Forcipomyia* sp. and *Atrichopogon* sp. were used as outgroups to root the tree.

Specimens identified based on wing patterns, genitalia, and head morphology as *C. leopoldoi*, *C. pusillus*, *C. ignacioi*, and *C. foxi* each clustered with their respective reference sequences in well-supported nodes, corroborating morphological assignments and supporting the utility of COI as a DNA barcode marker for species-level delimitation in these taxa. Specimens morphologically assigned to *C. venezuelensis* and additional taxa did not yield high-quality consensus sequences and were therefore excluded from phylogenetic reconstruction.

In contrast, morphotypes designated as *C. cf. insinuatus* and *C. cf. todatangae* grouped within the broader clade corresponding to the *C. paraensis* species complex and other members of the subgenus *Haematomyidium*, consistent with their diagnostic morphological affinity. However, no confidently identified reference sequences for these exact taxa were available for direct comparison. Additionally, *C. daviesi* formed a distinct lineage within the Mataemyia group, for which no other reference sequences were available in GenBank.

Pairwise COI genetic distances (K2P) further supported these phylogenetic patterns. Intraspecific divergence was low to moderate across taxa, including *C. foxi* (0.6–1.1%), *C. ignacioi* (0.3–2.0%), *C. insignis* (0.3–4.0%), *C. cf. insinuatus* (0.3–2.3%), *C. leopoldoi* (1.7%), and *C. pusillus* (2.3–4.3%). In contrast, interspecific divergence between *C. ignacioi* and the reference *C. guttatus* sequence was high (16–17%), supporting their molecular separation despite superficial morphological similarity. Distances within *Culicoides cf. todatangae* were low (0.6%), consistent with close relatedness between the two specimens. For the lineage assigned as *Culicoides daviesi* intraspecific divergence was 1.1%, whereas interspecific distances ranged from 16.9– 17.3% relative to *C. insinuatus*, 17–20% relative to *C. paraensis*, and 17–19% relative to *C. galindoi*.

### Haplotype diversity and genetic structure of *Culicoides insignis*

To further explore the genetic diversity of *Culicoides insignis*, the predominant species in Puerto Carreño, we analyzed the distribution of COI haplotypes obtained in this study together with previously reported sequences from other regions of the Americas. The network showed a compact topology, with most haplotypes separated by relatively few mutational steps and no apparent deep divergence among the sampled haplotypes (Figure 3).

**Figure 3.**
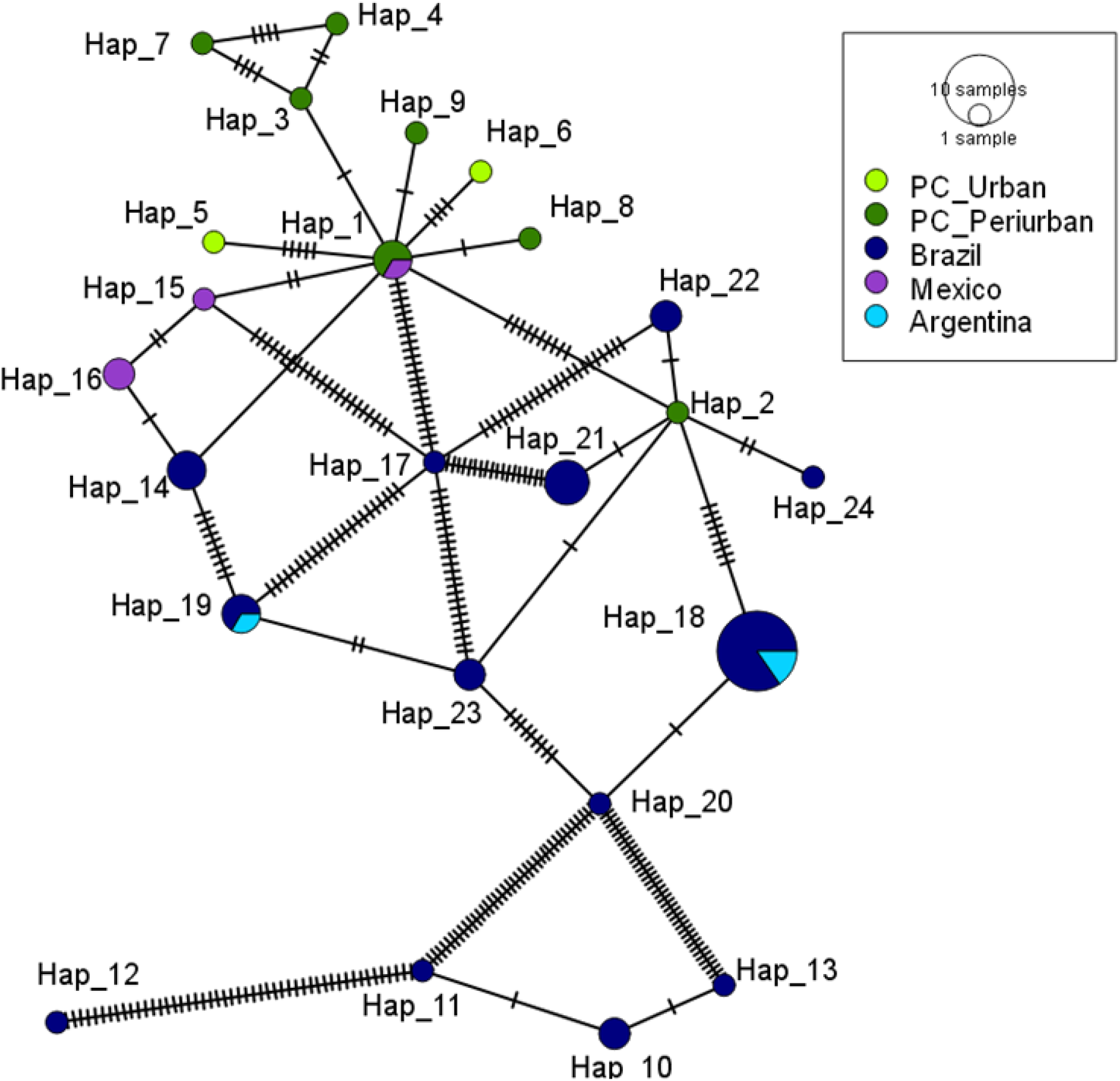
Minimum spanning haplotype network of *Culicoides insignis* inferred from a 379-bp fragment of the mitochondrial COI gene. The network illustrates genealogical relationships among COI haplotypes obtained in this study and previously reported from multiple regions of the Americas. Each circle represents a unique haplotype, with circle size proportional to its frequency. Colors denote geographic origin (Puerto Carreño and reference populations from Brazil, Argentina, and Mexico). Connecting lines represent mutational steps, and hatch marks indicate the number of nucleotide substitutions separating haplotypes.

Several haplotypes were shared between Puerto Carreño and populations from Brazil, Argentina, and Mexico. At the same time, exclusive haplotypes restricted to individual localities were detected. The network showed a compact topology, with most haplotypes separated by relatively few mutational steps and without evidence of deep phylogeographic structuring. Central nodes were broadly distributed across regions, whereas peripheral haplotypes accumulated additional substitutions and were less frequently represented.

### Blood-meal source analysis

A total of 64 blood-fed females were analyzed, including *C. insignis* (n = 56), C. ignacioi (n = 2), *C. pusillus* (n = 5), and *C. foxi* (n = 1). Three vertebrate blood-meal sources were identified: *Homo sapiens* (14% of analyzed specimens), *Equus caballus* (36%), and *Sus scrofa* (8%). These sources were identified in *C. insignis*, with horses being the most frequently detected host, followed by humans and pigs. Mixed blood meals involving humans and horses were also detected in six *C. insignis* specimens. Equine blood was additionally detected in *C. foxi* and *C. ignacioi*, whereas *C. pusillus* specimens showed evidence of feeding on humans and horses. These findings should be interpreted cautiously given the small number of specimens analyzed for species other than *C. insignis* (Table 3).

**Table 3.** Identification of the blood source for the analyzed *Culicoides* species.

| **Species** | ***Equus caballus*** | ***Homo sapiens*** | ***Sus scrofa*** | **Mixed** | **Total** |
|  | **Positive (%)** | **Positive (%)** | **Positive (%)** | **Positive\*(%)** | **Processed** |
|---|---|---|---|---|---|
| ***C. foxi*** | 1 (100) | - | - | - | 1 |
| ***C. ignacioi*** | 2 (100) | - | - | - | 2 |
| ***C. insignis*** | 19 (34) | 7 (12.5) | 5 (9) | 6 (10.7) | 56 |
| ***C. pusillus*** | 1(20) | 2(40) | - | - | 5 |
| **Total** | **23** (36) | **9** (14) | **5** (8) | **6** (9) | **64** |

\*The mixed component refers to organisms that were successfully amplified for two vertebrate hosts; in this case, all mixed interactions were exclusively associated with *H. sapiens* and *E. caballus*.

### Viral detection

The presence of OROV RNA was assessed by RT-qPCR targeting partial fragments of the L and S genomic segments in pooled specimens. The results are summarized in Table 4. *Culicoides insignis* showed the highest number of positive pools, with L segment amplification detected in 30 of 52 pools (57.7%) comprising 1,283 individuals. Accounting for unequal pool sizes, the estimated prevalence was 37.37 per 1,000 individuals (95% CI: 25.69–51.96). Detection using the S-segment assay was less frequent, with 7 of 44 pools (15.9%) testing positive among 1,111 individuals, corresponding to an estimated prevalence of 6.71 per 1,000 individuals (95% CI: 2.99–12.86). When a more conservative criterion requiring concordant detection of both genomic targets was applied, 3 of 32 pools were positive among 882 individuals, resulting in an estimated prevalence of 3.61 per 1,000 individuals (95% CI: 0.94–9.60). Interestingly, OROV RNA was detected in two females without visible blood remnants that also tested negative in blood-meal PCR assays. Thus, no evidence of a recent blood meal was detected in these specimens, and at least one positive *C. insignis* specimen was collected in an urban setting.

**Table 4.** Detection of oropuche virus by PCR using targets in the S and L segments in *Culicoides* spp.

| **Species** | **Molecular target** | **Positive/tested pools** | **Individuals tested** | **Estimated prevalence per 1,000 (95% CI)** |
|---|---|---|---|---|
| *C. insignis* | L | 30/52 | 1,283 | **37.37 (25.69–51.96)** |
|  | S | 7/44 | 1,111 | **6.71 (2.99–12.86)** |
|  | L + S* | 3/32 | 882 | **3.61 (0.94–9.60)** |
| *C. pusillus* | L | 4/6 | 24 | **371.01 (104.71–757.96)** |
|  | S | 0/3 | 19 | **0** |
|  | L + S* | 0/1 | 7 | **0** |
| *C. ignacioi* | L | 2/2 | 5 | **NE\*\*** |
|  | S | 1/4 | 12 | **87.13 (5.13–317.53)** |
|  | L + S* | 1/1 | 2 | **NE\*\*** |
| **Overall** | L | 36/60 | 1,312 | **44.76 (32.22–59.87)** |
|  | S | 8/51 | 1,142 | **7.45 (3.53–13.69)** |
|  | L + S* | 4/34 | 891 | **4.76 (1.54–11.17)** |

\*L + S represents concordant positivity for both molecular targets and includes only pools evaluated for both assays. \*\*NE, not estimable using the selected MLE/CI procedure because all evaluable pools were positive. Estimates based on small numbers of pools or individuals should be interpreted cautiously.

OROV L-segment RNA was also detected in *C. pusillus*, with four of six pools testing positive among 24 individuals, corresponding to an estimated prevalence of 371.01 per 1,000 individuals (95% CI: 104.71– 757.96). No S-segment amplification was detected in the three *C. pusillus* pools evaluated with this assay, and none of the pools evaluated for both targets showed concordant positivity. In *C. ignacioi*, both pools evaluated for the L segment were positive, representing five individuals; however, prevalence could not be reliably estimated because all evaluated pools were positive. One of four pools evaluated for the S segment was positive, corresponding to an estimated prevalence of 87.13 per 1,000 individuals (95% CI: 5.13–317.53). The only *C. ignacioi* pool evaluated for both targets was positive for both L and S, but the very small sample size precluded a reliable prevalence estimate. Because *C. pusillus* and *C. ignacioi* were represented by few individuals and pools, these estimates were highly uncertain and should not be interpreted as evidence of higher OROV prevalence than that observed in *C. insignis*.

### OROV sequencing

OROV-specific sequences were recovered from 13 pools, including nine *C. insignis*, three *C. pusillus*, and one *C. ignacioi*. Recovery of viral sequences was heterogeneous across samples and genomic segments, but reads mapping to the L, M, and S segments were detected in all 13 pools. Read-derived consensus sequences suitable for comparative analyses included 13 L-segment sequences spanning 173 nt, 11 M-segment sequences spanning 316 nt, and six S-segment sequences with up to 299 nt of homologous sequence. Longer overlapping M-segment sequences spanning 656 nt were additionally recovered from *C. insignis* (B76) and *C. pusillus* (B82) (Table S2).

Nucleotide variation among sequences was estimated using uncorrected p-distances. Comparisons among field-derived sequences and laboratory controls revealed nucleotide differences across the three segments. p-distances ranged from 0.01156 to 0.01734 for L (mean = 0.01445), from 0.00633 to 0.03165 for M (mean = 0.01251), and from 0.00334 to 0.02341 for S (mean = 0.01003). None of the field-derived sequences were identical to their corresponding laboratory controls across the analyzed regions.

The highest phylogenetic resolution was obtained from a 656-nt region of the M segment recovered from *C. insignis* (B76) and *C. pusillu*s (B82). The two field-derived sequences were identical across the complete analyzed region and differed from all 86 publicly available OROV sequences included in the analysis. Pairwise genetic distances were lower between B76/B82 and reference sequences assigned to the BR-2015–2024 lineage (mean p-distance = 0.00235; range = 0.00152–0.00457) than those assigned to the PE/CO/EC-2008– 2021 lineage (mean = 0.16599; range = 0.16463–0.16768). Consistent with these differences, Bayesian phylogenetic analysis grouped B76 and B82 with BR-2015–2024 sequences (Fig. 4). Complementary Bayesian analyses of shorter L (173 nt), M (316 nt), and S (299 nt) regions placed the remaining field-derived sequences within OROV genetic diversity, although these fragments provided limited resolution of relationships among closely related sequences (Fig. S2).

**Figure 4.**
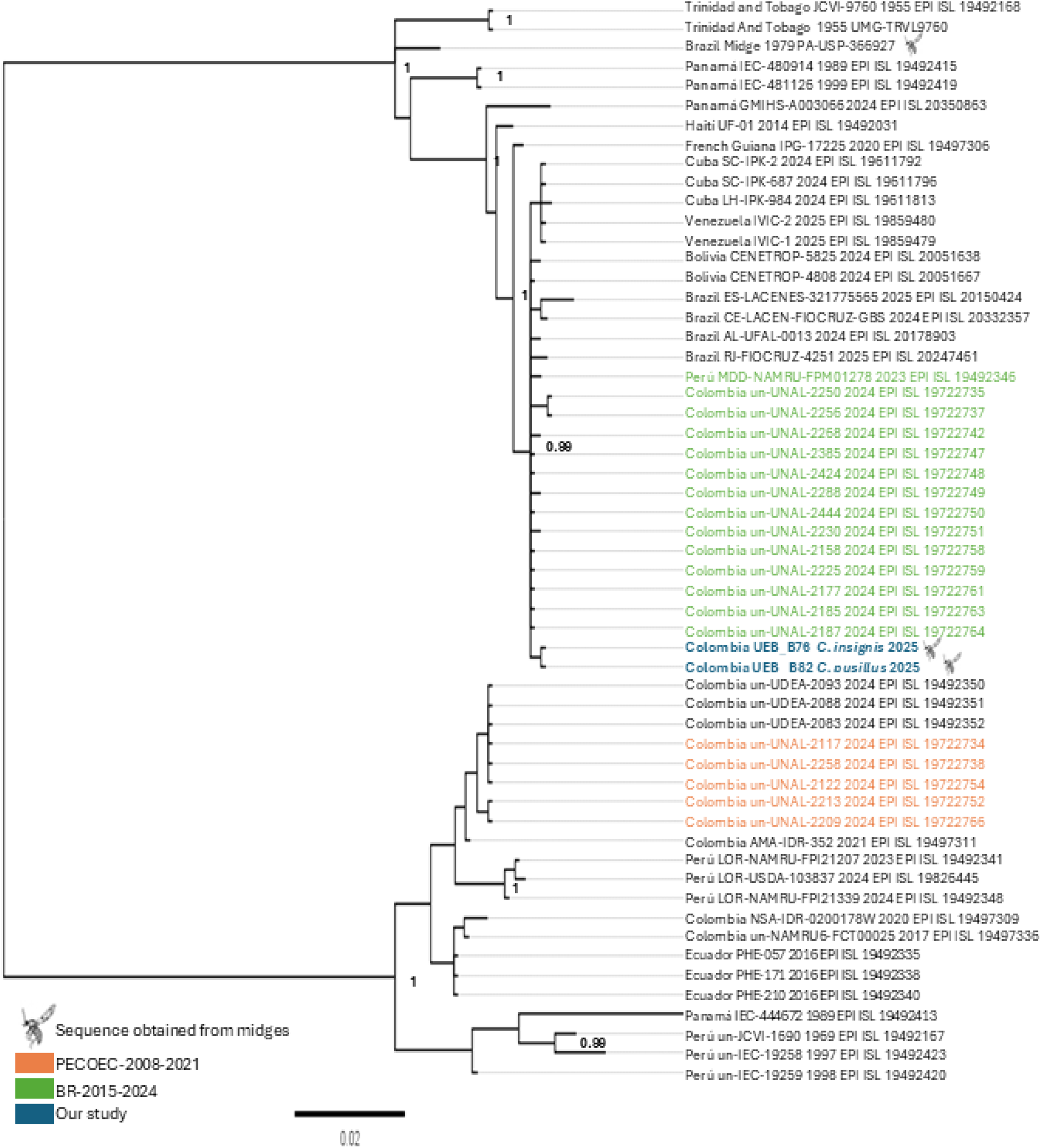
Bayesian phylogenetic analysis of the Oropouche virus M segment recovered from *Culicoides* spp. in Colombia. Bayesian phylogenetic reconstruction based on a 656-nt region of the OROV M segment, including sequences recovered from *Culicoides insignis* (B76) and *Culicoides pusillus* (B82) and publicly available OROV sequences. B76 and B82 are highlighted in blue. Reference sequences previously assigned to the BR-2015–2024 and PE/CO/EC-2008–2021 phylogenetic groups are shown in green and orange, respectively; sequences without a corresponding assignment in the published classification are shown in black. Values at nodes indicate Bayesian posterior probabilities **[≥0.99]**. The scale bar represents nucleotide substitutions per site.

The interaction network (Figure 5) integrates associations among *Culicoides* species, vertebrate blood-meal sources, and molecular evidence of OROV infection. Viral detection was based on RT-qPCR targeting the L and S segments and, in selected positive pools, was further supported by targeted Nanopore sequencing, which recovered OROV-specific sequences from all three genomic segments. *Culicoides* species with molecular evidence of OROV infection were associated with multiple vertebrate hosts, including humans, pigs, and horses. C*. insignis* showed the highest connectivity, reflecting its frequent viral detection and broader range of identified blood-meal sources, whereas *C. pusillus* and *C. ignacioi* showed fewer host associations. Overall, the network provides a descriptive integration of the virological and blood-meal findings across the three *Culicoides* species.

**Figure 5.**
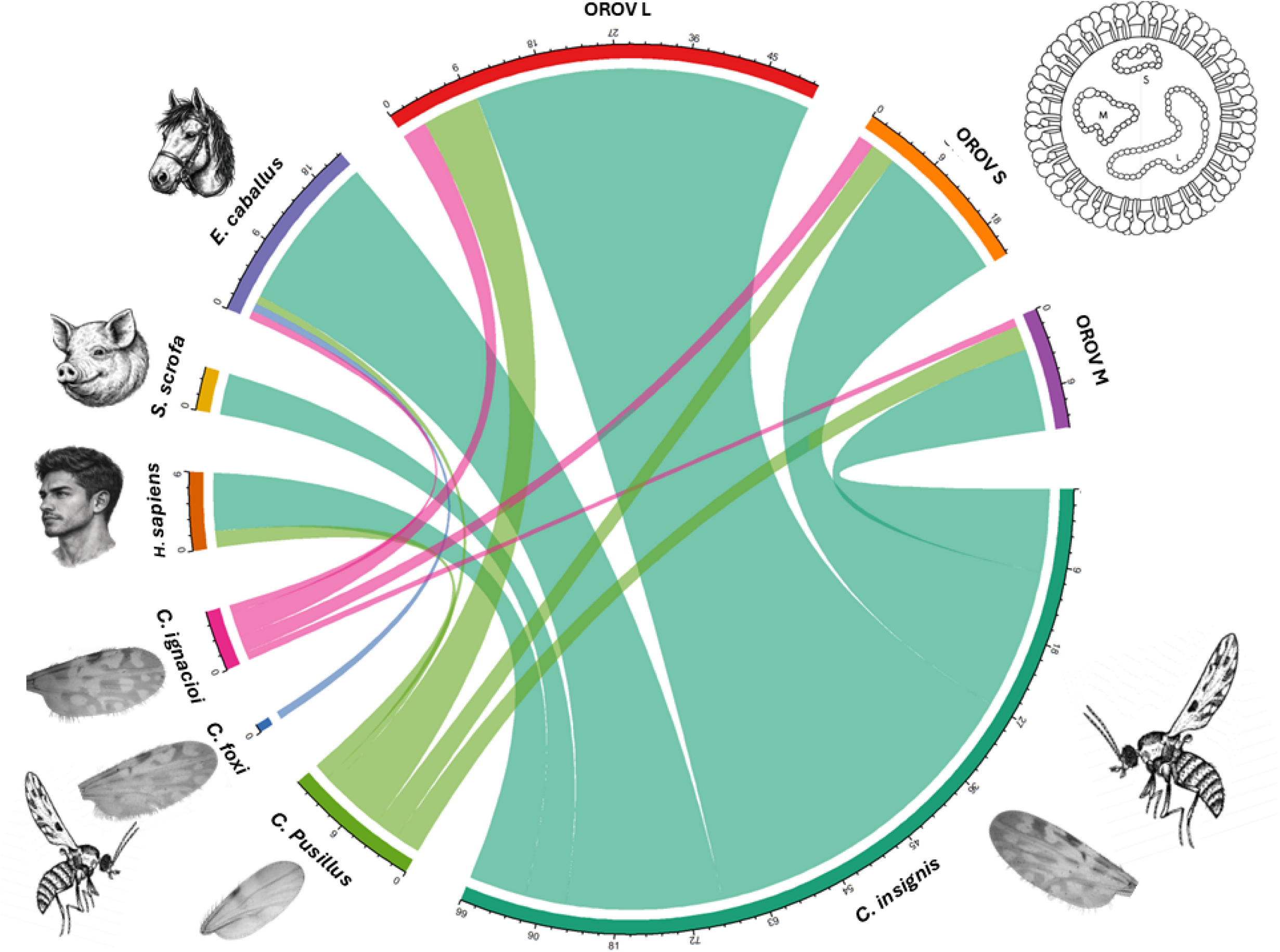
Chord diagram illustrating associations between *Culicoides* species, vertebrate blood-meal sources, and molecular detection of Oropouche virus RNA. The diagram provides a descriptive summary of blood-meal identification and viral detection results. Each colored chord corresponds to a *Culicoides* species and connects either to a vertebrate host identified through blood-meal analysis or to an OROV genomic segment detected by RT-qPCR or targeted Nanopore sequencing.

### Ecological analysis

Environmental predictors contributing to habitat suitability models differed among the four *Culicoides* species analyzed (Table 5, Figure S3), indicating species-specific differences in the environmental variables associated with predicted suitability. Although up to 12 predictors were retained after VIF filtering in several models, the number of predictors with positive drop-AUC values ranged from three in *C. ignacioi* to eight in both *C. paraensis* and *C. pusillus*, indicating differences in the number of independently informative environmental predictors across taxa. Model performance was moderate to high (AUC = 0.74–0.92; TSS = 0.56–0.68), with *C. pusillus* showing the strongest discrimination (AUC = 0.92; TSS = 0.68) (Table 5).

**Table 5.** Ecological niche model performance and drop-AUC–based variable importance across four *Culicoides* species.

| Species | Input |  |  | Model performance |  |  |  |  | Top five efective variables |  |  |  |  |
|---|---|---|---|---|---|---|---|---|---|---|---|---|---|
|  | **Presence data after cleaning (n)** | **Background points (n)** | **retained variables (n)** | **AUC** | **sensitivity** | **specificity** | **TSS** | **Effective variables (n)** | **Variable 1 (Drop AUC)** | **Variable 2 (Drop AUC)** | **Variable 3 (Drop AUC)** | **Variable 4 (Drop AUC)** | **Variable 5 (Drop AUC)** |
| *C. pusillus* | 83 | 1897 | 12 | 0.92 | 0.80 | 0.88 | 0.68 | 8 | Bio 03 (Isothermality) (0.03) | Elevation (masl) (0.01) | Bio 08 (0.006) | Slope (0.006) | Bio 19 (0.003) |
| *C. paraensis* | 59 | 1871 | 12 | 0.88 | 0.89 | 0.74 | 0.63 | 8 | Bio 03 (Isothermality) (0.06) | Bio 08 (0.004) | Bio 13 (0.001) | Bio 18 (0.006) | Bio10 (0.0005) |
| *C. insignis* | 83 | 1897 | 12 | 0.87 | 0.92 | 0.64 | 0.56 | 6 | Elevation (masl) (0.02) | Bio03 (Isothermality) (0.01) | Bio 18 (0.004) | Wind velocity (0.003) | Bio 10 (0.002) |
| *C. ignacioi* | 16 | 1110 | 6 | 0.74 | 0.80 | 0.79 | 0.59 | 3 | Bio07 (0.03) | Slope (0.002) | Bio15 (0.001) | 0 | 0 |

*C. ignacioi*, modeled with the smallest cleaned occurrence dataset (n = 16), exhibited three effective predictors, primarily annual temperature range (BIO07; drop-AUC = 0.03), slope, and precipitation seasonality (BIO15). Model performance was comparatively lower (AUC = 0.74; TSS = 0.59), and the limited number of occurrence records restrict further interpretation of its environmental associations.

*C. insignis* showed six effective predictors and moderate to high discrimination capacity (AUC = 0.87; TSS = 0.56). Elevation emerged as highest-ranked predictor (drop-AUC = 0.02), followed by isothermality (BIO03), precipitation during the warmest quarter (BIO18), wind speed, and BIO10, indicating contributions from both climatic and topographic variables (Table 5).

Although *C. paraensis* and *C. pusillus* both retained eight effective predictors, their ecological configurations differed. In *C. paraensis*, isothermality (BIO03; drop-AUC = 0.06) was the dominant contributor, with additional effects from precipitation metrics (BIO08, BIO13, BIO18) and soil-related variables, indicating contributions from temperature and precipitation-related variables. In contrast, *C. pusillus* displayed a more distributed contribution pattern, with BIO03 (drop-AUC = 0.03), elevation, BIO08, slope, and BIO19 contributing independently, indicating a different combination of informative environmental predictors (Table 5).

Pairwise Jaccard similarity indices based on retained effective predictors were generally low, indicating limited overlap in independently informative environmental variables among species. No shared effective predictors were detected between *C. ignacioi* and *C. insignis* (Jaccard = 0.00) or between *C. insignis* and *C. paraensis* (Jaccard = 0.00). The highest similarity was observed between *C. paraensis* and *C. pusillus* (Jaccard = 0.33), although substantial differentiation remained. Moderate similarity was observed between *C. ignacioi* and *C. pusillus* (Jaccard = 0.22), while other comparisons showed minimal overlap (≤ 0.10).

Overall, these results indicate differences in the environmental predictors contributing to model performance across species. Although temperature-related variables were recurrently informative, the composition of effective predictors, their drop-AUC values, and model discrimination metrics varied among taxa, supporting species-specific patterns in predicted environmental suitability.

### Co-occurrence analysis

Spatial patterns of predicted environmental suitability (threshold ≥ 0.4) varied among species and showed heterogeneous correspondence with the distribution of reported human OROV cases across the Americas (Figure 6). *C. paraensis* showed the clearest positive association between suitability and reported case occurrence, but without significant statistical association. In contrast, *C. insignis* displayed a broad and continuous pattern of suitability extending across northern South America, the Brazilian Atlantic coast, and Caribbean regions, including areas with limited or no reported case intensity. Predicted suitable areas extended beyond regions with historically documented OROV transmission. Suitability for *C. pusillus* and *C. ignacioi* showed partial overlap with regions reporting OROV cases in Brazil and the Amazon basin but also extended into areas with few or no reported cases. Overall, the maps indicate partial spatial concordance rather than strict geographic coincidence between environmental suitability and reported OROV cases, with substantial areas of predicted suitability occurring outside documented outbreak regions.

**Figure 6.**
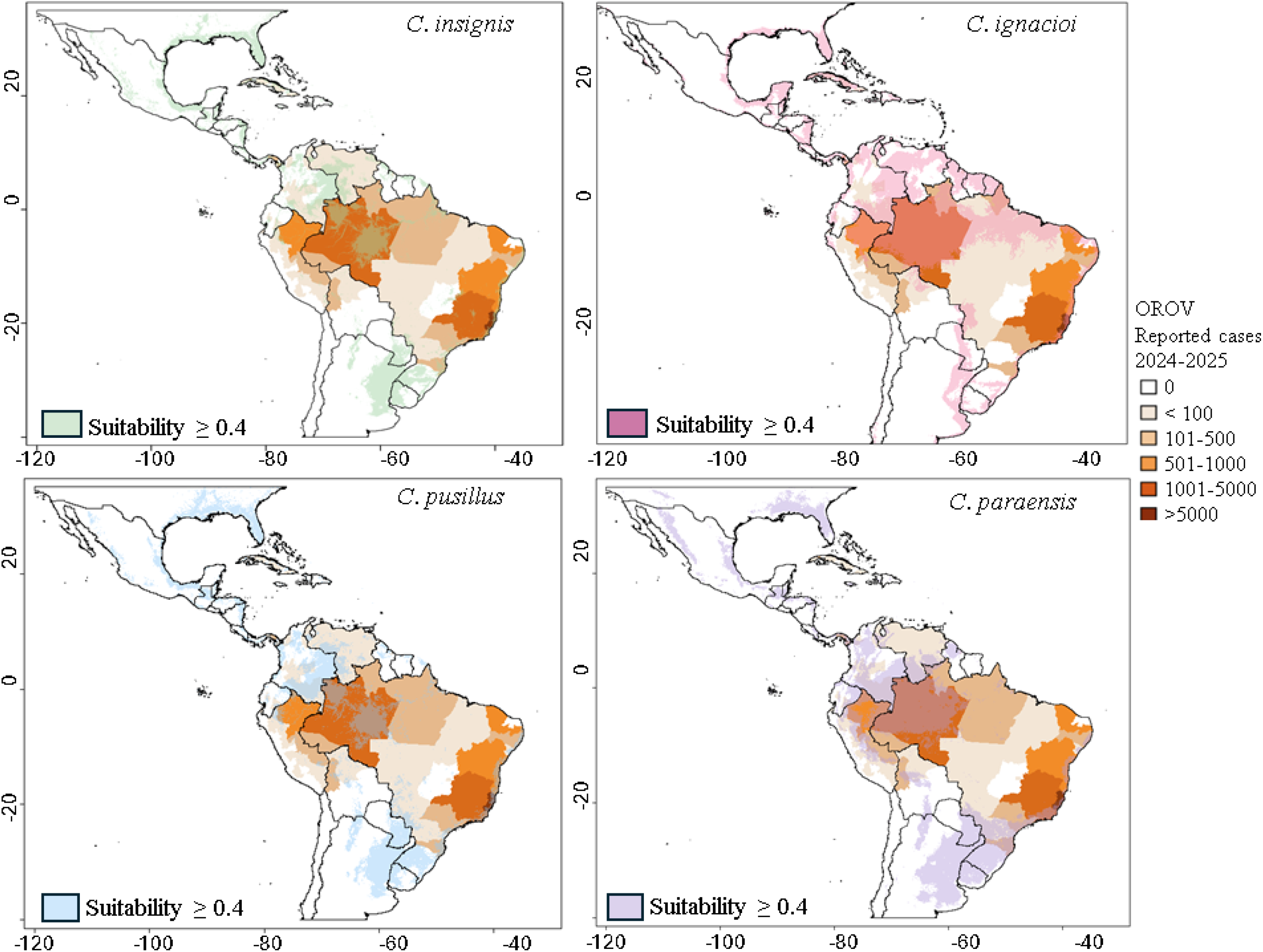
Ecological suitability of *Culicoides* species and spatial overlap with reported OROV cases in the Americas. Ecological niche models were generated using the maximum entropy algorithm (MaxEnt) to predict environmental suitability for *C. insignis*, *C. ignacioi*, *C. pusillus*, and *C. paraensis*. Areas with predicted suitability ≥ 0.4 are shown as species-specific colored overlays. Reported human Oropouche virus (OROV) cases for 2024–2025 were obtained from the PAHO/OPS and are grouped into five categories (≤100, 101–500, 501–1000, 1001–5000, and >5000). Country and regional boundaries were obtained from the Global Administrative Areas database (GADM), which provides spatial data for academic, non-commercial use and permits publication of derived maps under the Creative Commons Attribution license (CC-BY), in accordance with PLOS policies. All spatial analyses and map production were carried out in R.

To assess macro-scale ecological concordance between predicted suitability and reported OROV cases, mean suitability values were compared across first-level administrative units with and without confirmed cases. Among the four species evaluated, only *C. paraensis* showed the clearest positive association between suitability and reported case occurrence, although neither statistical test reached significance (Wilcoxon W = 6108, p = 0.059). Binary concordance analysis using a suitability threshold ≥ 0.4 indicated increased odds of case occurrence in suitable areas (OR = 2.03, 95% CI 0.74–5.05; Fisher’s exact p = 0.119), although this association did not reach statistical significance. Suitability distributions also differed between administrative units with and without reported cases for *C. ignacioi* (W = 10862, p = 0.046), but without significant binary concordance (OR = 0.64, 95% CI 0.27–1.52; p = 0.307). No evidence of concordance was detected for *C. pusillus* (W = 7838, p = 0.990; OR = 0.19, 95% CI 0.004–1.17; p = 0.104). For *C. insignis*, administrative units with suitability ≥ 0.4 did not overlap with reported case regions, resulting in a negative association in binary analysis (OR = 0, 95% CI 0–0.79; p = 0.023), although continuous suitability distributions did not differ significantly (W = 8148, p = 0.723). Overall, associations between predicted environmental suitability and reported human OROV cases varied among species and between analytical approaches. *C. paraensis* showed the most consistent direction of positive association, although statistical support was limited, whereas the remaining species showed weak, inconsistent, or negative associations at an administrative scale.

## Discussion

This study integrates taxonomic, trophic, virological, and ecological approaches to characterize *Culicoides* communities and their potential relevance to OROV ecology in an understudied region of Colombia. Morphological identification supported by COI barcoding documented local species diversity, while RT-qPCR and targeted sequencing provided molecular evidence of OROV RNA in *C. insignis*, *C. pusillus*, and *C. ignacioi*. Blood-meal analysis identified vertebrate hosts used by these species, and ecological niche models characterized their environmental associations across the Americas. Together, these findings provide species-specific evidence to guide entomovirological surveillance and prioritize further investigations into the potential involvement of *Culicoides* species in OROV transmission.

Applying an integrative taxonomic framework proved particularly informative for *Culicoides*, a genus characterized by subtle morphological differentiation and frequent cryptic species complexes. Reliance on a single line of evidence can lead to misidentifications and underestimation of diversity[17,47]. This study generated the first COI sequence dataset for *Culicoides* species from Colombia. Phylogenetic analyses revealed well-resolved clades for *C. insignis*, *C. ignacioi*, *C. foxi* and *C. pusillus*, largely consistent with morphological identifications based on wing, head, and genital characters. However, several morphotypes did not cluster with available reference sequences or form monophyletic groups, highlighting the need to report barcode sequences for neotropical *Culicoides* species, as their morphological and genetic diversity remains incompletely characterized. Species richness was lower than that reported in sylvatic and peridomestic areas of the Brazilian Amazon [34], but slightly higher than that documented in rural areas with livestock in Brazil, Peru, and Argentina in studies with comparable sampling approaches [48,49]. The community was strongly dominated by *Culicoides insignis*, accounting for over 70% of specimens and present at most sites. Similar dominance has been reported in Peru [49] and Argentina [50], supporting its characterization as synanthropic and ecologically tolerant. The marked intraspecific genetic variation observed in *C. insignis* is consistent with the diversity reported for widely distributed *Culicoides* species [51,52]. Although its geographic and population-level structure requires further investigation. The exclusive use of CDC light traps may have underrepresented *C. paraensis*, as human-attraction methods have shown higher collection efficiency for this species in other settings [53].

Molecular detection of OROV RNA in multiple *Culicoides* species provides evidence of the presence of the viral RNA in local midge populations. In *C. insignis*, prevalence estimates varied substantially according to the molecular criterion used, with the estimate based on concordant detection of both targets being substantially lower than those obtained from either molecular target individually. Using this more conservative criterion, the estimated prevalence was 3.61 per 1,000 individuals (95% CI: 0.94–9.60). Comparable entomological estimates for OROV remain scarce. During the 2024 outbreak in Cuba, OROV RNA was detected in one of two Ceratopogonidae pools analyzed, including 14 apparently non-engorged females in the positive pool, although specimens could not be identified to species [54]. In contrast, recent surveillance in outbreak areas of Minas Gerais, Brazil, detected no OROV-positive pools among 212 *C. paraensis* or 607 *C. leopoldoi* [53]. These contrasting findings emphasize the spatial and temporal heterogeneity of OROV detection in *Culicoides* populations and the limited availability of directly comparable entomological infection estimates. Interestingly, the prevalence estimated using concordant OROV detection in our *C. insignis* samples is of a similar order of magnitude to the 0.33% (approximately 3.3 per 1,000) BTV infection rate recently reported in unengorged *C. insignis* from Ecuador [55], although comparisons between different viruses, sampling designs, and analytical approaches should be interpreted cautiously. Furthermore, given the recognized role of *C. insignis* as a candidate vector of bluetongue virus in South America, the environmental conditions associated with its distribution in our models may also be relevant to surveillance of other *Culicoides* borne arboviruses, although their circulation including other Orthabunyavirus was not assessed in this study.

Although no OROV cases have been officially reported in Vichada, underdiagnosis or clinical misclassification with other febrile illnesses, such as dengue, cannot be excluded [4,56]. Detection of both L and S genomic segments in *C. insignis*, including some unfed females, suggests that viral RNA detection is unlikely to be solely associated with recent blood ingestion, although this does not demonstrate disseminated infection or vector competence. The marked difference in positivity between the L and S molecular targets highlights the importance of assay-dependent interpretation of OROV RNA detection. Differences in analytical sensitivity, target-region characteristics, or RNA integrity may have contributed to this discordance. Wise et al. [57] demonstrated that mismatches between diagnostic primers and an Ecuadorian OROV lineage compromised viral detection, with primer redesign improving analytical sensitivity by up to three orders of magnitude. This highlights how genetic variation in target regions can affect molecular detection and should be considered when interpreting differences between diagnostic targets.

Sequence-based characterization provides an additional level of molecular support for OROV detection in *Culicoides* and allows these detections to be interpreted within the genetic diversity currently recognized in South America. Recent genomic surveillance has revealed substantial OROV diversity in South America, including the co-circulation and changing relative frequency of genetically distinct viral populations in the Colombian Amazon [23,58,59]. In this context, the phylogenetic proximity of the midge-derived M-segment sequences to sequences associated with the recent outbreak is consistent with contemporary regional OROV diversity. Importantly, however, the field-derived sequences were not identical to previously reported sequences across the homologous regions analyzed; a pattern observed both for the longer M-segment region and for most of the shorter genomic fragments. This differentiation may reflect previously unsampled OROV viral diversity, particularly because OROV had not previously been documented or genetically characterized in Vichada, although the limited genomic coverage prevents determining whether these differences represent geographically structured diversity or broader variation among circulating OROV populations.

Recent genomic studies have demonstrated that the evolutionary relationships of OROV may differ among its three genomic segments, and formal assignment of contemporary lineages has relied on complete or near-complete genomic information [23,58,59]. Therefore, the partial sequences recovered here should not be interpreted as evidence for formal lineage assignment or reassortment. Instead, they support the genetic characterization of OROV detected in field-collected *Culicoides* and highlight the need to obtain complete or near-complete viral genomes directly from biting midges to determine how OROV diversity in vectors relates to that characterized from contemporary human infections in Colombia and elsewhere in South America.

Blood meal analysis revealed opportunistic feeding by *C. insignis* on horses (*Equus caballus*), humans (*Homo sapiens*), and pigs (*Sus scrofa*), consistent with findings from Ecuador[55]. Similar feeding patterns in other hematophagous dipterans in Colombia suggest that shared vertebrate hosts in anthropogenically modified landscapes may facilitate viral spillover [60]. Although horses are not currently considered competent amplifying hosts for OROV, serological surveys in Brazil have reported neutralizing or hemagglutination-inhibiting antibodies against orthobunyaviruses in equines [61,62], indicating exposure in peri-urban and rural settings.

A central contribution of this study is the comparative ecological characterization of potential epidemiological interest *Culicoides* species. Previous niche modeling of *C. paraensis* emphasized temperature-related variables and precipitation seasonality as dominant drivers of distribution [12]. Our models recover part of this signal, particularly temperature stability (BIO03) and precipitation metrics. Prior modeling of *C. insignis* identified temperature, precipitation during the warmest quarter, elevation, and vegetation indices as key predictors [63,64]. Our results are broadly concordant, with elevation, precipitation variables (BIO18, BIO19), and wind speed emerging as independently informative predictors. Vegetation and soil variables exerted comparatively minor effects. Although precipitation did not uniformly dominate model performance, it creates temporary changes in surface moisture, and larval habitat availability [50]. Recent models have further identified humidity, soil moisture, and agricultural land use, particularly banana cultivation, as important correlates of *C. paraensis* suitability, consistent with the use of decaying banana stems and cocoa residues as larval habitats, while phylogeographic analyses have associated banana and cocoa cultivation, human population density, and *C. paraensis* suitability with OROV expansion [65,66].

Environmental predictors identified among *Culicoides* species in our study, suggest that their ecological associations may be more heterogeneous than those commonly represented in mosquito-centered arbovirus models, highlighting the need to consider multiple vector species and their species-specific habitat requirements. Wind-driven movement is increasingly recognized as facilitating range expansion and connectivity in *Culicoides* [67], providing a plausible ecological explanation for the broad geographic distribution and genetic connectivity observed in *C. insignis,* although its role in shaping the distribution of this species in the study region remains untested. Furthermore, as occurrence records were not spatially thinned, geographic clustering and uneven sampling intensity in biodiversity databases may have influenced model predictions and performance metrics. Accordingly, the ecological suitability estimates should be interpreted as exploratory, particularly for species represented by relatively few occurrence records.

Spatial comparisons revealed heterogeneous associations between *Culicoides* predicted suitability and reported OROV cases. Previous projections for *C. paraensis* have shown discrepancies with reported Oropouche fever occurrence, interpreted as potential underdetection or involvement of additional vectors [12]. This heterogeneity likely reflects the coarse spatial resolution of available epidemiological data and within-unit environmental heterogeneity, where focal high-suitability areas may be diluted when averaged across large administrative regions. Stronger concordance for *C. paraensis* aligns with its recognized epidemiological role. Given the broad environmental suitability predicted for multiple potential epidemiological interest *Culicoides* species, including *C. paraensis*, the geographic extent of ecological permissiveness may exceed the currently documented distribution of human cases. OROV infection presents nonspecific febrile symptoms overlapping with dengue and other arboviral illnesses, increasing the likelihood of underdiagnosis and misclassification in passive surveillance systems [5]. Moreover, discrepancies were observed between ecological suitability and reported incidence [68]. Furthermore, reported case occurrence is influenced by surveillance intensity, diagnostic capacity, population density, human mobility, healthcare access, and reporting practices, none of which were explicitly considered in our spatial comparisons; therefore, ecological concordance should not be interpreted as direct evidence of epidemiological relevance or vector involvement.

Taken together, the taxonomic, trophic, virological, and ecological findings support further investigation of *Culicoides* species other than *C. paraensis* in the context of OROV circulation in Colombia. Among the species examined, *C. insignis* warrants particular attention because of its local abundance, repeated molecular detections of OROV, including in pools of females without detectable blood meals, and evidence of feeding on humans and domestic animals. OROV-derived sequences were also recovered from *C. pusillus* and *C. ignacioi*, although their limited sample sizes and less extensive host-feeding data restrict further interpretation of their epidemiological relevance. These findings do not establish additional OROV vectors or demonstrate a multi-vector transmission system. Rather, they identify species for which experimental studies of viral infection, dissemination, and transmission are needed to clarify their potential roles in OROV ecology.

In conclusion, integrating taxonomy, ecology, host-feeding patterns, and virological evidence provides a practical approach to identifying and prioritizing *Culicoide*s species for further investigation in understudied regions. By generating convergent evidence within the same surveillance framework, this approach moves beyond isolated viral detections and single-species assumptions and enables candidate vectors to be progressively prioritized according to multiple complementary criteria. Although these findings do not establish additional OROV vectors, they illustrate how integrative entomovirological surveillance can simultaneously detect emerging viruses, characterize their ecological context, and generate the evidence needed to direct vector-competence studies and subsequent vector-incrimination efforts.

## Data Availability

All data supporting this study are available in the manuscript and its Supporting Information files. COI and Oropouche virus (OROV) sequences generated in this study were deposited in NCBI GenBank, with accession numbers provided in Tables S1 and S2, respectively. Pool-level RT-qPCR results and Nanopore sequencing metrics are included in Table S2. Multiple sequence alignments used for phylogenetic reconstruction are provided as supplementary ZIP files. Occurrence records used for ecological niche modeling were obtained from GBIF, published datasets, and field collections, as described in the Methods. Model performance results and predicted suitability maps are presented in Table 5 and Figure S3.

## ACKNOWLEDGEMENTS

We thank the administrative staff of the Secretaría de Salud del Vichada, particularly Dr. Jorge Luis de las Salas Ali, and Dr. Katheryn Barrios, for their support. We are thankful for the cooperation of Dr. Johanna Madroñero with the initial filtration and curation of the nanopore sequences corresponding to OROV. We are especially grateful to our field, laboratory collaborators, Jorge Galeano, and Emilio Bocanegra. We also thank the local communities from urban and rural areas of Puerto Carreño for receiving us and allowing us to conduct this research. Finally, we thank Dr. Ligia Moncada for her valuable comments and recommendations, and Dr. Maria Clara Alves from Oswaldo Cruz Foundation, for helping us to clarify *Culicoides daviesi* taxonomical determination.

## Supplementary figures

**Figure S1.**
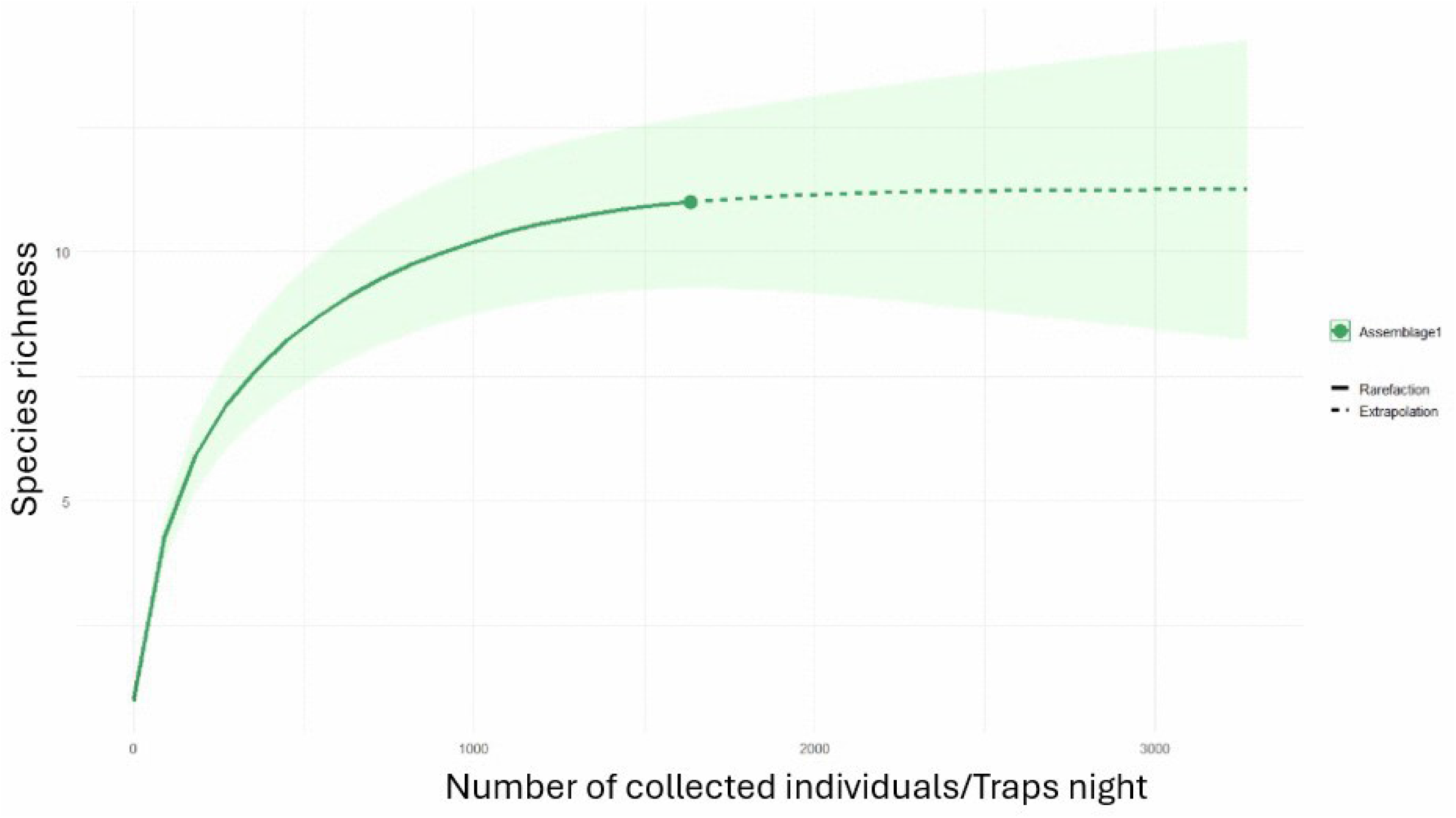
Rarefaction curve of *Culicoides* species richness as a function of sampling effort (traps per night).

**Figure S2.**
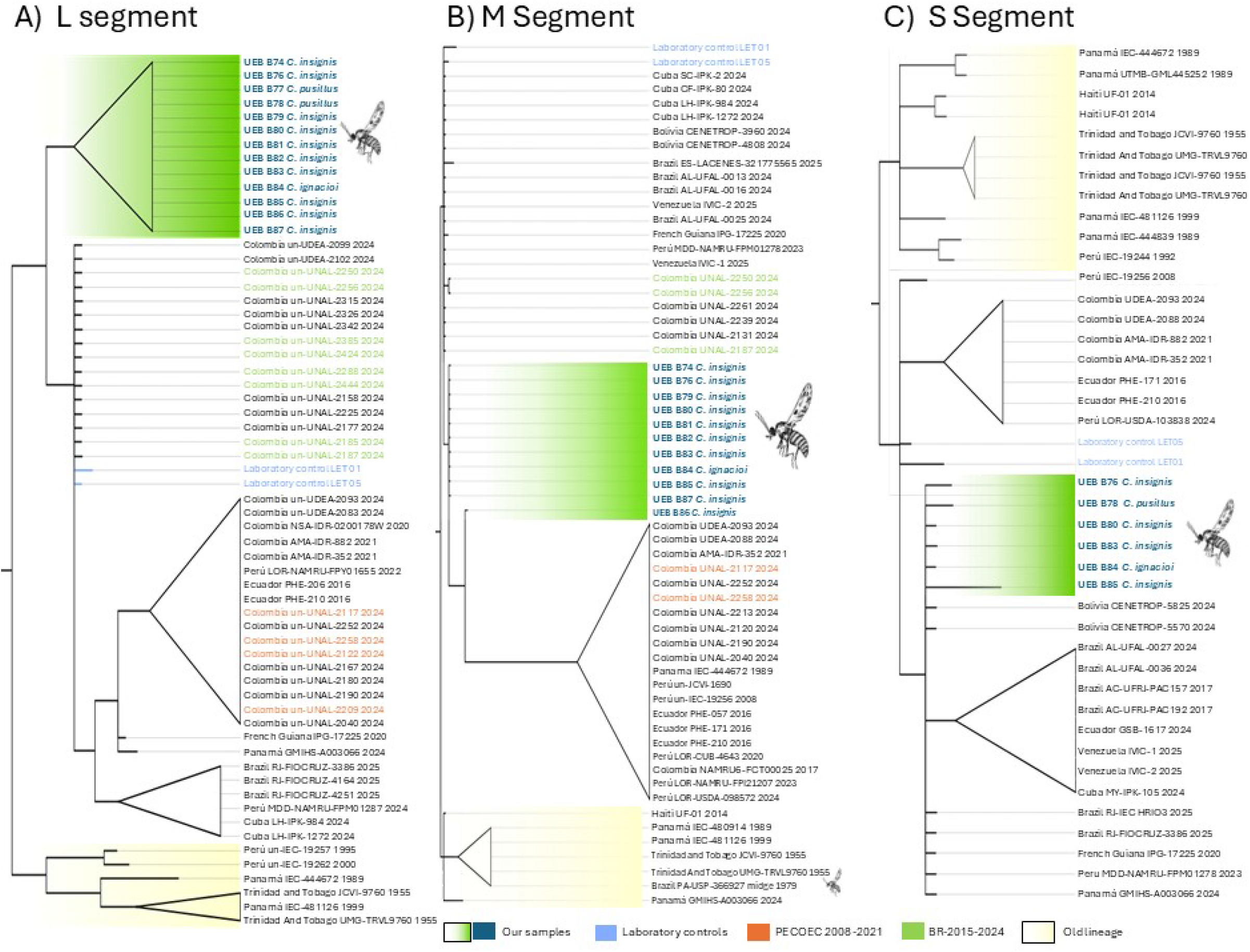
Bayesian phylogenetic analyses of partial OROV genomic sequences from the L, M, and S segments. Bayesian phylogenetic trees inferred from partial sequences of the A) L (173 nt), B) M (316 nt), and C) S (299 nt) genomic segments of Oropouche virus (OROV). Field-derived sequences obtained from *Culicoides* specimens were compared with homologous sequences available in GenBank. Separate Bayesian phylogenetic analyses were performed for each genomic segment using the corresponding aligned regions. The trees show the phylogenetic relationships between field-derived sequences and previously reported OROV sequences.

**Figure S3.**
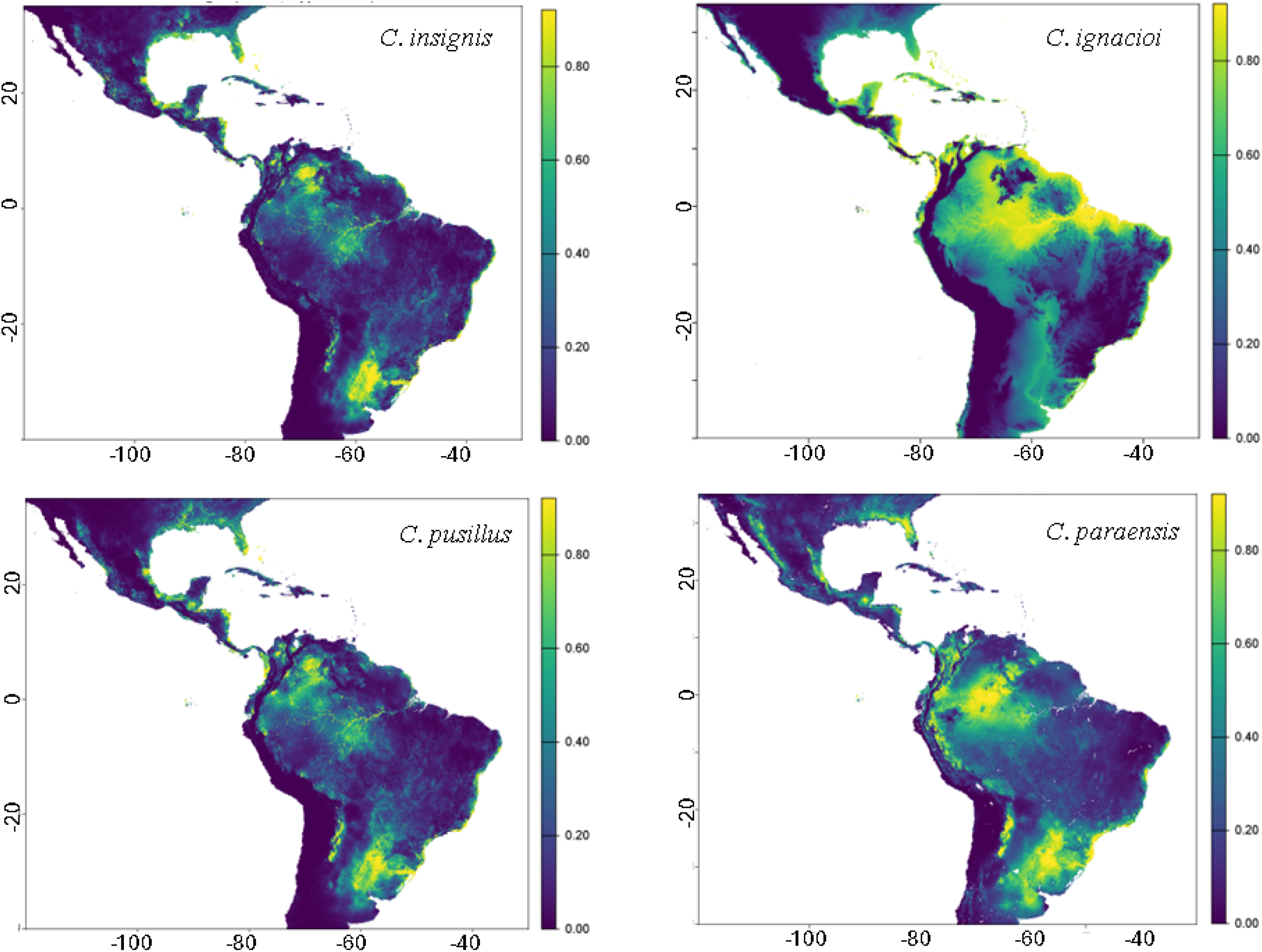
Potential distribution modeling of *Culicoides* species using MaxEnt. Predicted suitability values range from low (purple) to high (yellow), indicating the likelihood of environmentally suitable conditions for *C. insignis, C. ignacioi, C. pusillus, and C. paraensis* across the Americas.

**Table S1.**
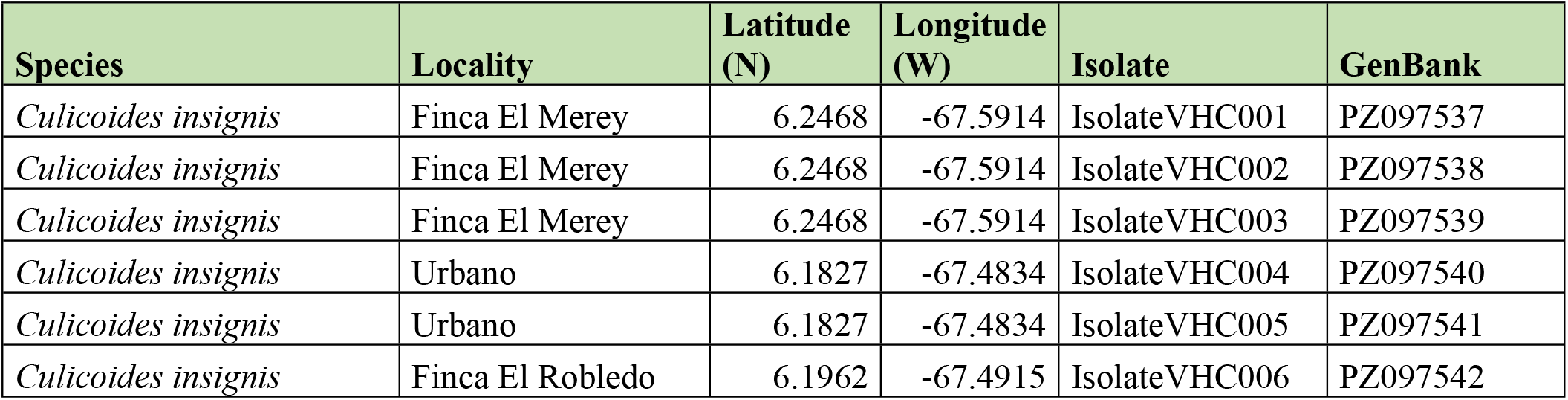
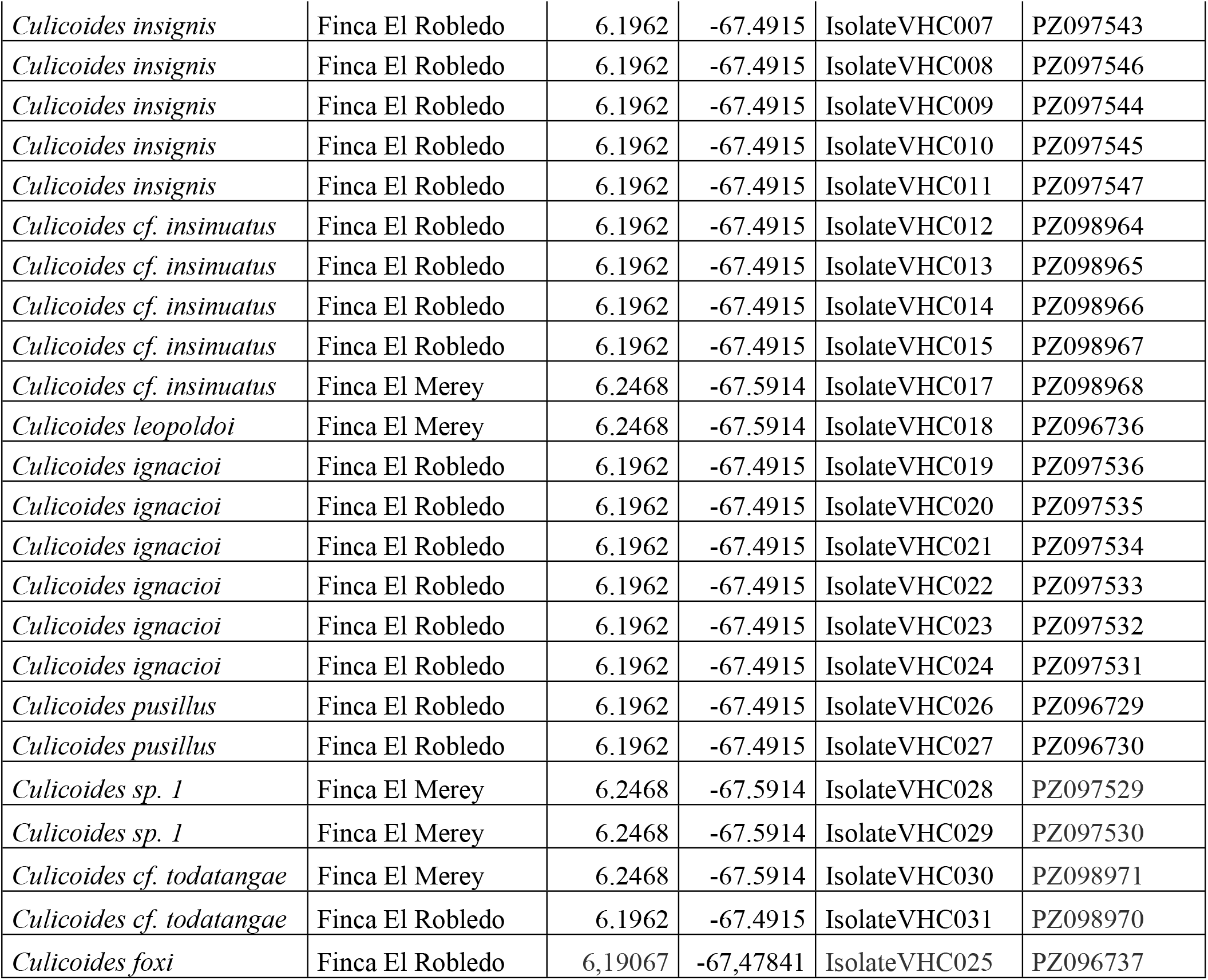
Samples description, collection locality, and GenBank accession numbers assigned.

| **Species** | **Locality** | **Latitude (N)** | **Longitude (W)** | **Isolate** | **GenBank** |
|---|---|---|---|---|---|
| *Culicoides insignis* | Finca El Merey | 6.2468 | -67.5914 | IsolateVHC001 | PZ097537 |
| *Culicoides insignis* | Finca El Merey | 6.2468 | -67.5914 | IsolateVHC002 | PZ097538 |
| *Culicoides insignis* | Finca El Merey | 6.2468 | -67.5914 | IsolateVHC003 | PZ097539 |
| *Culicoides insignis* | Urbano | 6.1827 | -67.4834 | IsolateVHC004 | PZ097540 |
| *Culicoides insignis* | Urbano | 6.1827 | -67.4834 | IsolateVHC005 | PZ097541 |
| *Culicoides insignis* | Finca El Robledo | 6.1962 | -67.4915 | IsolateVHC006 | PZ097542 |
| *Culicoides insignis* | Finca El Robledo | 6.1962 | -67.4915 | IsolateVHC007 | PZ097543 |
| *Culicoides insignis* | Finca El Robledo | 6.1962 | -67.4915 | IsolateVHC008 | PZ097546 |
| *Culicoides insignis* | Finca El Robledo | 6.1962 | -67.4915 | IsolateVHC009 | PZ097544 |
| *Culicoides insignis* | Finca El Robledo | 6.1962 | -67.4915 | IsolateVHC010 | PZ097545 |
| *Culicoides insignis* | Finca El Robledo | 6.1962 | -67.4915 | IsolateVHC011 | PZ097547 |
| *Culicoides cf. insinuatus* | Finca El Robledo | 6.1962 | -67.4915 | IsolateVHC012 | PZ098964 |
| *Culicoides cf. insinuatus* | Finca El Robledo | 6.1962 | -67.4915 | IsolateVHC013 | PZ098965 |
| *Culicoides cf. insinuatus* | Finca El Robledo | 6.1962 | -67.4915 | IsolateVHC014 | PZ098966 |
| *Culicoides cf. insinuatus* | Finca El Robledo | 6.1962 | -67.4915 | IsolateVHC015 | PZ098967 |
| *Culicoides cf. insinuatus* | Finca El Merey | 6.2468 | -67.5914 | IsolateVHC017 | PZ098968 |
| *Culicoides leopoldoi* | Finca El Merey | 6.2468 | -67.5914 | IsolateVHC018 | PZ096736 |
| *Culicoides ignacioi* | Finca El Robledo | 6.1962 | -67.4915 | IsolateVHC019 | PZ097536 |
| *Culicoides ignacioi* | Finca El Robledo | 6.1962 | -67.4915 | IsolateVHC020 | PZ097535 |
| *Culicoides ignacioi* | Finca El Robledo | 6.1962 | -67.4915 | IsolateVHC021 | PZ097534 |
| *Culicoides ignacioi* | Finca El Robledo | 6.1962 | -67.4915 | IsolateVHC022 | PZ097533 |
| *Culicoides ignacioi* | Finca El Robledo | 6.1962 | -67.4915 | IsolateVHC023 | PZ097532 |
| *Culicoides ignacioi* | Finca El Robledo | 6.1962 | -67.4915 | IsolateVHC024 | PZ097531 |
| *Culicoides pusillus* | Finca El Robledo | 6.1962 | -67.4915 | IsolateVHC026 | PZ096729 |
| *Culicoides pusillus* | Finca El Robledo | 6.1962 | -67.4915 | IsolateVHC027 | PZ096730 |
| *Culicoides sp. 1* | Finca El Merey | 6.2468 | -67.5914 | IsolateVHC028 | PZ097529 |
| *Culicoides sp. 1* | Finca El Merey | 6.2468 | -67.5914 | IsolateVHC029 | PZ097530 |
| *Culicoides cf. todatangae* | Finca El Merey | 6.2468 | -67.5914 | IsolateVHC030 | PZ098971 |
| *Culicoides cf. todatangae* | Finca El Robledo | 6.1962 | -67.4915 | IsolateVHC031 | PZ098970 |
| *Culicoides foxi* | Finca El Robledo | 6,19067 | -67,47841 | IsolateVHC025 | PZ096737 |

**Table S2.** OROV detection by RT-qPCR and Nanopore-derived sequence characterization in *Culicoides* specimens.

